# Computer-aided interpretation of chest radiography reveals the spectrum of tuberculosis in rural South Africa

**DOI:** 10.1101/2020.09.04.20188045

**Authors:** Jana Fehr, Stefan Konigorski, Stephen Olivier, Resign Gunda, Ashmika Surujdeen, Dickman Gareta, Theresa Smit, Kathy Baisley, Sashen Moodley, Yumna Moosa, Willem Hanekom, Olivier Koole, Thumbi Ndung’u, Deenan Pillay, Alison D. Grant, Mark J. Siedner, Christoph Lippert, Emily B. Wong, the Vukuzazi Team

## Abstract

Computer-aided digital chest radiograph interpretation (CAD) can facilitate high-throughput screening for tuberculosis (TB), but its use in population-based active case finding programs has been limited. In an HIV-endemic area in rural South Africa, we used a CAD-algorithm (CAD4TBv5) to interpret digital chest x-rays (CXR) as part of a mobile health screening effort. Participants with TB symptoms or CAD4TBv5 score above the triaging threshold were referred for microbiological sputum assessment. During an initial pilot phase, a low CAD4TBv5 triaging threshold of 25 was selected to maximize TB case finding. We report the performance of CAD4TBv5 in screening 9,914 participants, 99 (1.0%) of whom were found to have microbiologically proven TB. CAD4TBv5 was able to identify TB cases at the same sensitivity but lower specificity as a blinded radiologist, whereas the next generation of the algorithm (CAD4TBv6) achieved comparable sensitivity and specificity to the radiologist. The CXRs of people with microbiologically-confirmed TB spanned a range of lung field abnormality, including 19 (19.2%) cases deemed normal by the radiologist. HIV-serostatus did not impact CAD4TB’s performance. Notably, 78.8% of the TB cases identified during this population-based survey were asymptomatic and therefore triaged for sputum collection on the basis of CAD4TBv5 score alone. While CAD4TBv6 has the potential to replace radiologists for triaging CXRs in TB prevalence surveys, population-specific piloting is necessary to set the appropriate triaging thresholds. Further work on image analysis strategies is needed to identify radiologically-subtle active TB.

## Introduction

Tuberculosis (TB) continues to cause over 1 million deaths annually, challenging the WHO strategy to reduce deaths due to TB by 95% by 2035.^1^ Efficient strategies to find the “missing millions” are of particularly high priority in resource-limited settings where TB burden is high and access to diagnostics may be limited.^2,3^ In this context, community-based screening programs may play an important role in increasing case finding.^4–6^ However, these programs are challenged by high costs of TB diagnostic tests, including molecular tests such as Xpert MTB/RIF® and microbiological culture.^7,8,9^ The WHO recommends using chest radiography to select individuals with lung field abnormalities for sputum testing.^8^ However, this approach requires a workforce of radiologically-trained clinicians to interpret chest x-rays (CXR), which is often limited in rural low-resource settings.^11^

Computer-aided detection (CAD) systems have the potential to make TB screening programs more efficient by rapidly detecting TB-related abnormalities in digital CXRs. In December 2020, the WHO announced that forthcoming recommendations for systematic screening for TB will include the deployment of CAD algorithms.^12^ One commercial product, CAD4TB (©Thirona, Nijmegen, the Netherlands) scores CXRs on a scale from 0-100 according to degree of abnormality and the likelihood of TB.^13,14^ Previous studies have shown that CAD4TB can identify CXRs with abnormal lung fields and TB-related features, such as cavities and consolidation,^15,16–18^ and reduce the cost for microbiologic diagnostics.^14,19^ Using CAD for triage requires selecting a decision threshold score above which participants are referred for sputum testing. Although a universally applicable threshold would be useful, several studies have suggested that threshold selection should be adjusted according to factors unique to each setting, including underlying TB prevalence, demographic parameters, type and number of available microbiological tests.^13,20,21^ Another consideration that may impact computer-assisted CXR-interpretation is that HIV-positive patients may show atypical radiological signs of TB.^22,23^ So far, CAD4TB has been mainly evaluated in clinical settings in which symptomatic patients seek diagnostic evaluation. One study evaluated CAD4TB version 5 (CAD4TBv5) retrospectively at a diabetes-care center in Indonesia and found triaging threshold 65 had 88.9% sensitivity.^24^ Performance reports from population-based screening programs of asymptomatic individuals are limited. One study applied CAD4TBv5 retrospectively in a population-based setting and suggested lowering the CAD4TB score threshold for triaging compared to a clinical setting.^25^ How best to determine thresholds in prospective applications for active case-finding remains an unanswered question.

Here we report the prospective application of CAD4TBv5 as a triaging tool during a community-based multi-morbidity survey in a TB- and HIV-endemic area in rural South Africa.^26^ Mobile health camps were used to provide free health assessment to local residents over the age of 15 years, including measurement of body mass index (BMI), blood-pressure and glycosylated hemoglobin (HbA1c) and HIV-status. The goal of the TB component of the survey was to identify as many active TB cases as possible and to describe the full spectrum of TB disease in the population (Supplementary Figure 1). Following WHO-guidelines^10^, we aimed to triage participants for sputum testing if they reported TB-related symptoms or if they had abnormal lung fields on CXR. Because it was infeasible to employ a radiologist to review CXRs in real-time in the mobile health camp, we applied CAD4TBv5 to indicate lung field abnormality as triaging criterion for sputum collection. Sputum was subjected to Xpert Ultra MTB/RIF (Xpert Ultra) and liquid mycobacterial culture. A senior radiologist, who was blinded to CAD4TB output and other participant data, interpreted each CXR in a central reading setting and assessed whether a) lung fields were normal or abnormal, and b) if abnormal, whether the findings were diagnostic of active TB. Participants who were determined to have abnormal lung fields by the radiologist but whose CAD4TB scores were below the triaging threshold were visited at home by the research team to collect a sputum sample for microbiological assessment.

Here we report the process by which we selected a triaging threshold to maximize the number of participants with abnormal lung fields that were triaged for sputum collection at the mobile health camp and the performance of CAD4TBv5 (and its successor CAD4TBv6) to identify active TB on CXRs obtained in a community-based prevalence survey.

## Results

### Selection of a CAD4TBv5 triaging threshold

During a pre-planned pilot phase of the survey, 1,090 participants underwent chest radiography and were triaged for sputum examination based on a CAD4TBv5 triage threshold of 60, a threshold that had been chosen to be more than 80% sensitive for active TB based on existing literature at the time.^17,24^ At this threshold, CAD4TBv5 identified only 54 of 198 (27.3%) CXRs which the radiologist subsequently determined to have abnormal lung fields. Participants were considered incorrectly triaged if they were ultimately found to have lung field abnormality by the radiologist, but had been sent home without sputum collected at the mobile health camp. 109 (83.2%) of the incorrectly triaged participants (n=131) eventually had successful sputum collection at home, and 22 (16.8%) of incorrectly triaged participants could not be contacted for follow-up visit and therefore did not have a sputum collected. The high rate of incorrect triaging posed logistical challenges to the operation of the survey. Using the radiologist’s assessment of lung field abnormality as gold standard, the range of CAD4TBv5 scores was between 10-100 for normal (median 23, interquartile range (IQR) 20-28) and overlapped with those for abnormal lung fields (range 17-100, median 43, IQR 30-61) (Supplementary Figure 2). In order to minimize the number of participants that were incorrectly triaged in the field and maximize TB case-finding, a triaging threshold of 25 was determined to be optimal for the main phase of the study. At this threshold the sensitivity for detecting lung field abnormality was as high as possible (84.8% (95% confidence interval (CI): 79.1-89.5) given the trade-off in specificity (65.7, CI: 62.5-68.8, Supplementary Figure 3), which had to account for the increased burden on the laboratory to perform sputum molecular and microbiological testing.

### Characteristics of participants with TB

A total of 10,320 participants were enrolled in the screening program (Figure 1). 406 participants could not have chest radiography due to pregnancy or physical inability to climb into the mobile van; they are excluded from this analysis of CAD4TB performance. Based on their CXR findings and microbiology results, 9,914 participants were categorized as having 1) “definite TB”, defined by microbiological evidence of *Mycobacterium tuberculosis* (Mtb) in the sputum (either positive MTB/RIF XpertUltra or Mtb culture); 2) “probable TB”, defined as radiological findings consistent with active TB without any microbiological evidence of TB; or 3) “no evidence of active TB”, defined by absence of radiological and microbiological evidence of TB. The median age of participants in the survey was 39 years (interquartile range (IQR) 24-59), 29.8% (2,954 of 9,914) were HIV-positive and 10.7% (1,056 of 9,914) presented with at least one TB-related symptom. Among the 99 participants (1.0%) who met the definition for definite TB, only 20 (20.2%) reported any TB symptom (Table 1). According to the radiologist’s assessment of the CXRs from this group, 30 (30.3%) had lung field abnormalities diagnostic of TB and 80 (80.8%) had any lung field abnormality. The median CAD4TBv5 score of this group was 63 (IQR 44-77.5, range 19-100).

**Figure 1:**
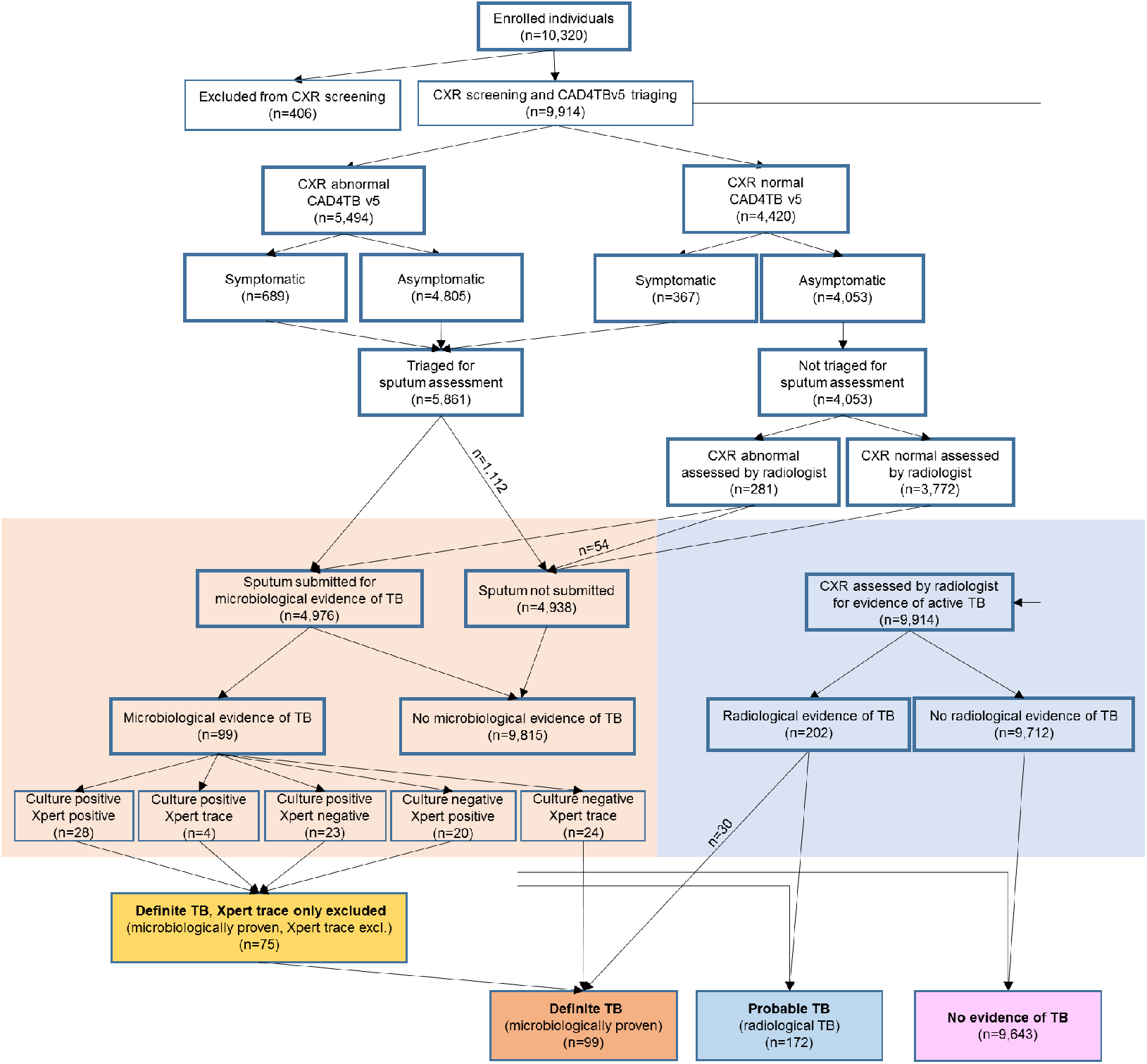
TB screening in the first year of Vukuzazi. A total of 10,320 participants were enrolled, of which 9,914 underwent chest radiography. Participants were triaged for sputum collection if they had symptoms or if their CAD4TBv5 score was equal to or above the triaging threshold. For triaged participants who could spontaneously produce a sputum sample (n=4,976), sputum was tested with Xpert MTB/RIF Ultra and liquid culture. Independently, the radiologist indicated whether chest radiographs were normal or abnormal, and if abnormal were diagnostic of active TB. If the radiologist indicated abnormal lung fields among participants who were not triaged in the camp, sputum was collected during a home-visit. Participants were grouped into definite (microbiologically-confirmed), probable (radiologically diagnosed), and no evidence of TB. A more stringent sub-group of definite TB excluded participants whose only microbiological evidence of TB was a Xpert Ultra “trace” result.

**Table 1:** Demographics, clinical and radiological characteristics of study participants. Characteristics are listed among all enrolled participants who underwent chest radiography (n=9,914) and by TB diagnostic group: definite TB (microbiologically-confirmed), definite TB, trace excluded (more stringent group that excludes those whose only microbiological evidence of TB is a Xpert “trace” results), probable TB (radiologically diagnosed). Characteristics are shown as absolute frequencies (relative frequencies in %), or median* (25%-75% interquartile range (IQR)) for age and CAD4TB scores.

| **Characteristic** | **Enrolled individuals with CXR** | **Definite TB** | **Definite TB trace excluded** | **Probable TB** | **No TB** |
| --- | --- | --- | --- | --- | --- |
| N | 9,914 (100) | 99 (1.0%) | 75 (0.8%) | 172 (1.7) | 9,643 (97.3) |
| Female | 6,664 (67.2) | 49 (49.5) | 35 (46.7) | 76 (44.2) | 6,539 (67.8) |
| Age* | 39 (24-59) | 51 (31-63.5) | 50 (31.5-63.5) | 56 (41.8-63.3) | 38 (23-58) |
| HIV positive | 2,954 (29.8) | 41 (41.4) | 34 (45.3) | 76 (44.2) | 2,837 (29.4) |
| On ART (of HIV positive) | 2,408 (81.5) | 34 (82.9) | 28 (82.4) | 65 (85.5) | 2,309 (81.4) |
| History of TB | 1,196 (12.1) | 25 (25.3) | 17 (22.7) | 110 (64.0) | 1,061 (11.0) |
| Current smoker | 726 (7.3) | 17 (17.2) | 14 (18.7) | 30 (17.4) | 679 (7.0) |
| Cough currently | 677 (6.8) | 14 (14.1) | 12 (16.0) | 39 (22.7) | 624 (6.5) |
| Fever | 295 (3.0) | 9 (9.1) | 7 (9.3) | 13 (7.6) | 273 (2.8) |
| Weight loss in prior 6 months | 232 (2.3) | 10 (10.1) | 8 (10.7) | 10 (5.8) | 212 (2.2) |
| Night sweat | 326 (3.3) | 8 (8.1) | 6 (8.0) | 12 (7.0) | 306 (3.2) |
| Any TB symptom | 1,056 (10.7) | 20 (20.2) | 15 (20.0) | 45 (26.2) | 991 (10.3) |
| Any lung field abnormality according to radiologist | 2,002 (20.2) | 80 (80.8) | 64 (85.3) | 171 (99.4) | 1,751 (18.2) |
| Chest x-ray diagnostic of TB according to radiologist | 202 (2.0) | 30 (30.3) | 25 (33.3) | 172 (100) | 0 (0) |
| CAD4TBv5 scores* | 28 (22-41) | 63 (44-77.5) | 66 (51.5-84) | 87 (73-95) | 27 (22-40) |
| CAD4TBv6 scores* | 35 (16-46) | 61 (48.5-78) | 64 (52-78) | 78 (68-87) | 34 (16-46) |

The wide spectrum of lung field abnormality and CAD4TBv5 scores observed among participants with definite TB is illustrated by two examples from the extremes of the spectrum; the first with prominent radiological abnormalities deemed by the radiologist to be consistent with active TB and by CAD4TBv5 as highly consistent with TB with score of 95 (Figure 2a-b) and the second with lung fields deemed normal by the radiologist and by CAD4TBv5 as having a low likelihood of active TB with a score of 39 (Figure 2c-d). Notably, both these participants reported being asymptomatic and were therefore triaged for sputum collection based on their CAD4TBv5 scores alone.

**Figure 2:**
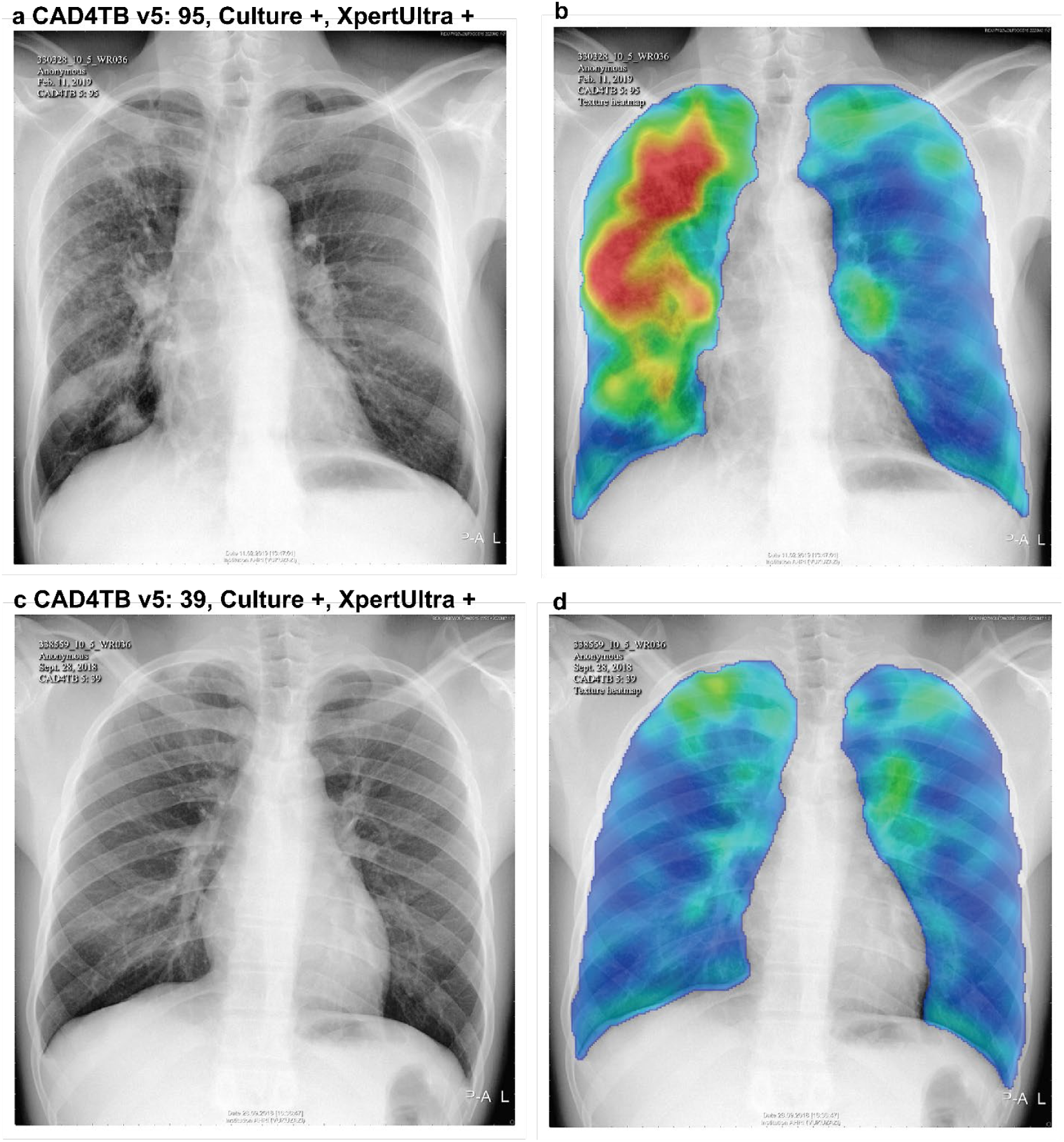
Radiographic spectrum of two participants with microbiologically-confirmed active TB. Digital CXR images and corresponding CAD4TBv5 heatmaps (blue colors mark normal lung fields and red marks TB-related abnormalities contributing to CAD4TB score). Participant 1 (a, b) was 62 years old, HIV-positive on ART and had a CAD4TB v5 score of 95 which the radiologist assessed as abnormal and diagnostic of TB. Participant 2 (c, d) was 31 years old, HIV-negative and had a CAD4TB v5 score of 39 which the radiologist assessed as normal. Both participants had positive liquid culture and XpertUltra (greater than trace) results, no TB symptoms, and no history of previous TB treatment.

Among the participants with definite TB, 24 had a XpertUltra “trace” result (lowest level in the semiquantiative scale^27^) as the only microbiological evidence of TB. These participants were excluded from the more stringent sub-group, “definite TB, trace only excluded” (75/9,914, 0.76%, Figure 1) which was defined to allow sensitivity analyses due to emerging uncertainty about the clinical significance of the XpertUltra “trace” laboratory result when present in isolation.^28^ The median CAD4TBv5 score for this group was 66 (IQR 51.5-84) and did not differ significantly from the definite TB group (median 63 (IQR 43-78), p-value=0.43). An additional 172 (1.7%) participants met the definition of “probable TB” and the median CAD4TBv5 score in this group was significantly higher (87 (IQR 73-95), p-value=5.3×10^−11^) than in the definite TB group. In individuals with definite and probable TB, there were no significant differences for either version of CAD4TB when groups were stratified by HIV-status (p-values >0.05, Figure 3).

**Figure 3:**
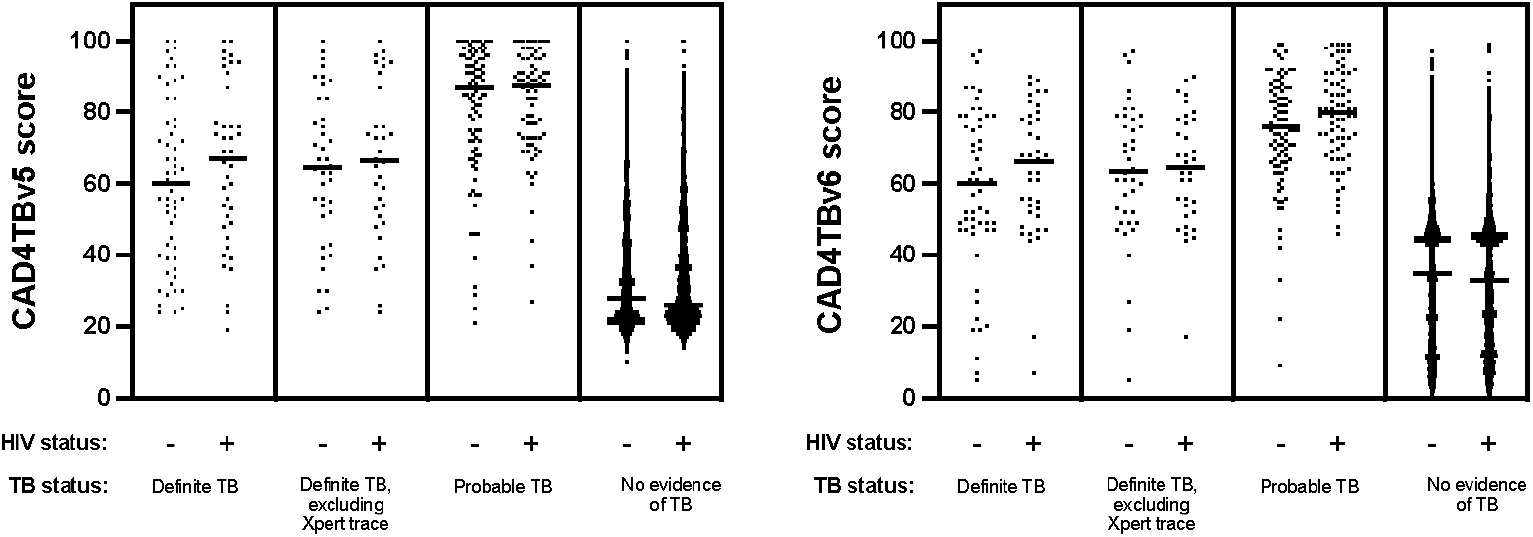
CAD4TB scores differ by TB but not HIV status. CAD4TB v5 (a) and v6 (b) distributions across TB diagnostic groups: definite TB (microbiologically-proven n=99), definite TB, trace excluded (more stringent group that excludes those whose only microbiological evidence of TB is a Xpert “trace” results, n=75), probable TB (radiologically diagnosed, n=172), and no evidence of TB (n=9,643). Scores in each group were stratified by HIV status (definite TB HIV-positive: n=41, HIV-negative: n=57; definite TB, trace excluded HIV-positive: n=34, HIV-negative: n=40; probable TB HIV positive: n=76, HIV-negative: n=96; no TB HIV-positive: n=2,837, HIV-negative: 6,755). Horizontal lines mark the median. P-values from comparisons between HIV-positive vs. HIV-negative in each diagnostic group were for definite TB: 0.26, definite TB, trace excluded: 0.70, probable TB: 0.88, no TB 2.9×10^−5^.

### Performance of CAD4TB as a tool to triage participants for sputum examination

We assessed the performance of CAD4TBv5 and v6 to correctly triage participants with definite TB compared to the expert radiologist’s performance (Supplementary Tables 1, 2, and 3). The radiologist’s triage sensitivity for definite TB (80.8%, CI: 71.7-88.0), was most closely comparable to CAD4TBv5’s performance between the scores of 39 and 40 (39: 82.8% (CI: 73.9-89.7), 40: 79.8% (CI: 70.5-87.2) and CAD4TBv6’s performance between the scores of 47 and 48 (47: 82.8% (CI:73.9-89.7), 48: 76.8% (CI: 67.2-84.7), Table 1). At these thresholds the specificity of CAD4TBv5 (39: 55.4 (CI: 54.0-56.8), 40: 57.4% (CI: 56.0-58.8)) was lower than that of the radiologist (66.9 (CI: 65.6-68.2)). The specificity of CAD4TBv6 at these thresholds was higher (47: 62.6 (CI: 61.2-64), 48: 68.0 (CI: 66.7-69.3)) and in the same range as the radiologist. At these thresholds, CAD4TBv5 also had lower (39: 3.6% (CI: 2.9-4.5), 40: 3.7% (CI: 2.9-4.5)) and CAD4TBv6 had similar precision (positive predictive value) (47: 4.3% (CI: 3.4-5.3), 48: 4.6% (CI: 3.7-5.8)) when compared to the radiologist (4.7% (CI: 3.8-5.8)).

CAD4TB’s triage performance to detect the more stringent sub-group (definite TB, trace excluded) did not differ significantly from its performance to detect definite TB (v5: AUC 0.82 (0.77-0.87) vs. AUC 0.78 (CI: 0.73-0.83); v6: 0.84 (0.79-0.89) vs. 0.79 (0.73-0.84), p-value=0.28, Fig 4a-b, Supplementary Table 4). However, CAD4TB’s triage performance for the detection of probable TB (for both v5 and v6: AUC 0.96, CI: 0.95-0.98) was superior to its performance for the detection of definite TB (v5: AUC 0.78 (CI: 0.73-0.83), v6: AUC 0.79 (CI: 0.73-0.84), p-values <0.001 in Supplementary Table 5, Fig 4a-b). The performance of CAD4TB did not differ significantly when participants were stratified by HIV status (Supplementary Table 6). Similarly, the area under the precision-recall curve (PRAUC) was lowest for definite TB trace excluded (both v5 and v6: 0.08) and definite TB (both v5 and v6: 0.09) and highest for probable TB (v5: 0.42, v6: 0.44) (v5: Supplementary Figure 4a and v6: Supplementary Figure 4b).

**Figure 4:**
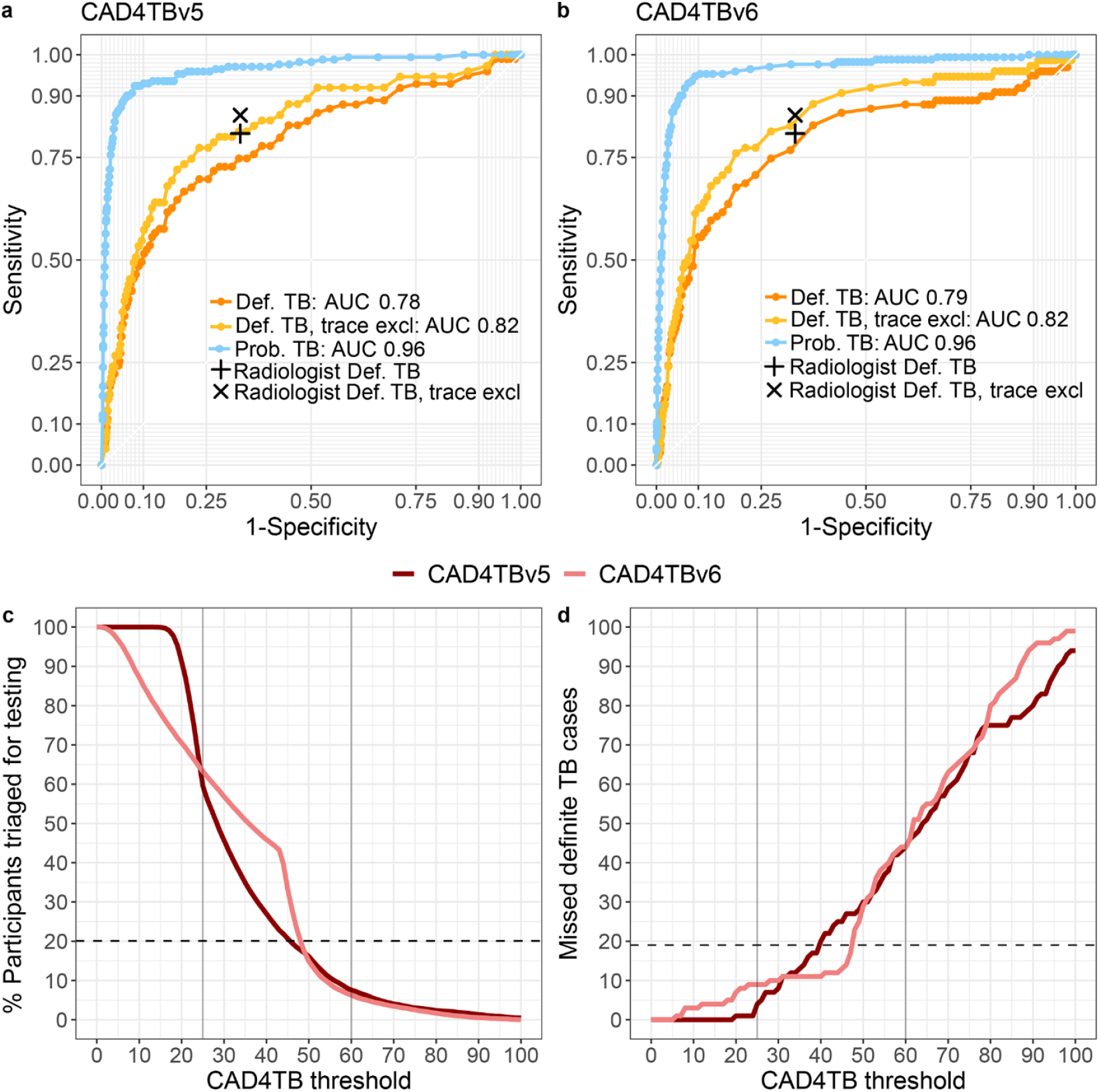
Performance of CAD4TB v5 and v6. Receiver operating curve of CAD4TBv5 (a) and v6 (b) triaging thresholds for each TB diagnostic group: definite TB (microbiologically-confirmed, positive: n=99, negative: n=4,877), definite TB, trace excluded (more stringent group that excludes those whose only microbiological evidence of TB is a Xpert “trace” results, positive: n=75, negative: 4,901), probable TB (radiologically diagnosed, positive: n=172, negative: n=9,742). The radiologist’s sensitivity and specificity of detecting definite TB (+) and definite TB trace excluded (x) is marked. The (c) percentage of participants who would have required sputum testing at each CAD4TB triaging threshold (n=9,914) and (d) number of definite TB cases that would have been missed at each CAD4TB triaging threshold (n=99). In (c) and (d) the horizontal dashed line indicates the radiologist’s performance and vertical lines indicate the CAD4TBv5 triaging thresholds used in the pilot (60) and main study phase (25).

If the radiologist’s definition of abnormal lung fields had been used for field triage, 20.2% (2,002) of participants would have required sputum testing, (Table 2, Figure 4c). At triaging thresholds that matched the radiologist’s sensitivity, CAD4TBv5’s would have required testing a larger percentage of participants (39: 28.4% (2,820), 40: 27.0% (2,677)) and CAD4TBv6 would have required testing a percentage of participants similar to the radiologist (47: 23.7% (2,348), 48: 20.1% (1,997), Table 2 and Figure 4c). The radiologist’s number needed to test to identify one person with definite TB (NNT) was 25, which was lower compared to CAD4TBv5 (39: 34, 40: 34) and similar to CAD4TBv6 (47: 29, 48: 26, Supplementary Figure 5). Our study used an intentionally low CAD4TBv5 triaging threshold of 25, which prompted sputum collection for the majority of all study participants (5,906 (59.6%), Figure 1) and identified 19 definite TB cases (19.2% of all definite TB cases) that the radiologist assessed as having ‘normal lung fields’. These cases would have been missed if we had not used such a low triaging threshold (Table 2, Figure 4d). Notably, only four of these cases reported TB symptoms (which would have resulted in sputum collection), therefore 15 of these individuals were asymptomatic and would have remained undetected by radiologist triage. Both versions of CAD4TB would have missed a similar number of definite TB cases (v5: 39: 17 (17.2%) 40: 20 (20.2%); v6: 47: 17 (17.2%), 48: 23 (23.2%)). Four definite TB cases had CAD4TBv5 scores below the triaging threshold of 25 and were triaged for sputum because the participants screened positive for TB symptoms.

**Table 2:**
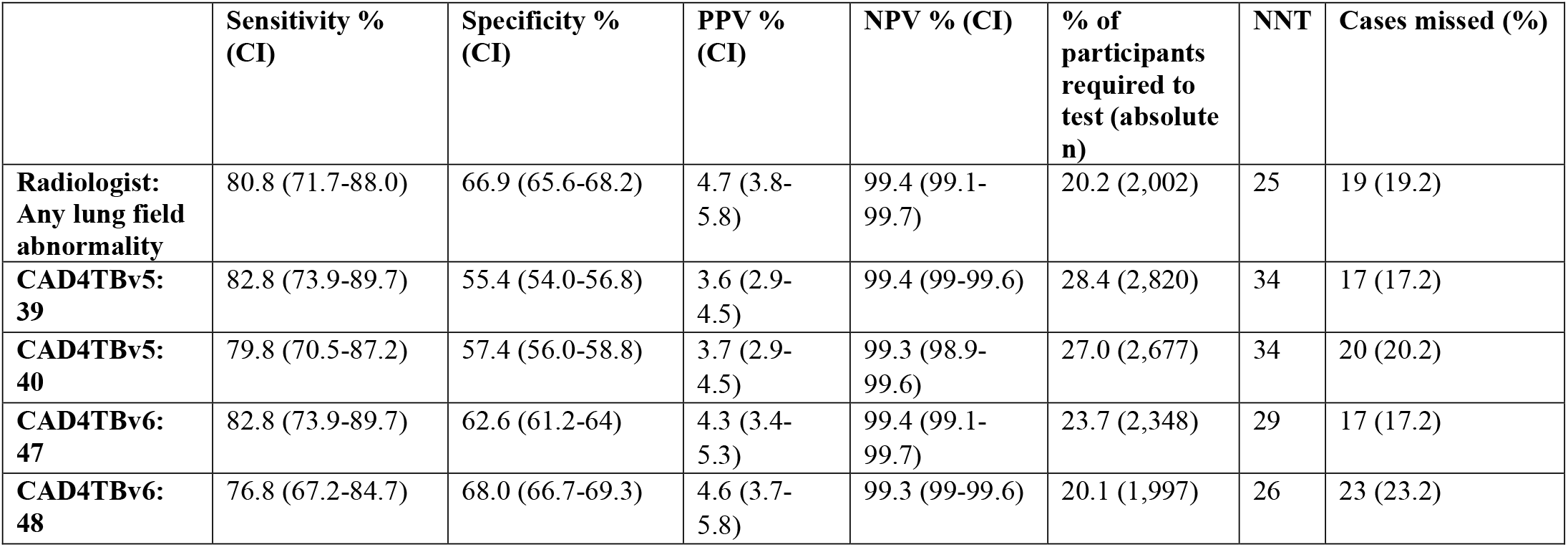
Performance of CAD4TBv5 and v6 to identify definite TB at the radiologist’s sensitivity. Definite TB was defined as microbiological evidence, either by positive XpertUltra or culture. Presented are the radiologist’s sensitivity to triage definite TB by classifying ‘any lung field abnormality’ in comparison to CAD4TBv5 and v6 scores that closed matched the radiologist’s sensitivity. Performance is given as sensitivity, specificity, positive predictive value (PPV), negative predictive value (NPV) with 95% confidence intervals (CI); percentage of participants required to be tested at this threshold, the number of needed to test (NNT) to identify one person with definite TB and the number of missed definite TB cases.

## Discussion

We used CAD4TBv5 to interpret digital CXRs in a community-based multi-disease screening survey in rural South Africa and found that CAD4TB could perform similarly to an expert radiologist in triaging participants for diagnostic sputum testing. At a triaging threshold 39, CAD4TBv5 had a similar sensitivity but slightly lower specificity than the radiologist; however, retrospective analysis of the next generation of the algorithm, CAD4TBv6, showed that a triaging threshold of 47 had equivalent sensitivity and improved specificity, making its performance comparable to that of the radiologist. At a triaging threshold of 47, CAD4TBv6 would have triaged slightly more participants (346) for sputum testing and identified 2 more TB cases than the radiologist.

The importance of screening for TB in community-based non-clinical settings is increasingly recognized. Our study, which used mobile health camps in a rural area, found that 1.0% of the screened population had microbiologically-proven TB cases, and that 79.8% of these individuals were asymptomatic. This finding is in line with a recent study from a population-based screening in urban Uganda, which showed that individuals newly diagnosed with active TB in community-based screening have fewer symptoms, compared to people diagnosed at health facilities.^29^ It is also in line with the recently completed South African National TB prevalence survey (57.7% asymptomatic)^30^ and a recent meta-analysis of global TB prevalence surveys (79-97% asymptomatic)^31^. Asymptomatic TB eludes traditional symptom-based TB screening algorithms but is identifiable by CXR abnormality in most cases.^30,31^ Because of this, the WHO has recently announced that upcoming TB screening guidelines will recommend a symptom-agnostic community-based approach using CXR surveys.^12^ Our results suggest that using CAD4TBv6 to triage digital CXRs will increase the yield of active case finding programs compared to symptom-based triage and is feasible in rural low-resourced settings. The truck containing the radiography unit for our study was equipped with solar panels that powered the x-ray equipment and computer, allowing it to be used without external sources of electricity. The off-line CAD4TB algorithm interpreted CXRs for triaging within one minute of their capture without requiring an active internet connection. The numerical CAD4TB read-out made it straightforward for non-clinical staff to appropriately direct participants for sputum collection. Portable digital radiography units and CAD algorithms that perform comparably to an expert radiologist make large-scale CXR screening feasible and relevant to find the missing millions of undiagnosed TB.^32^

In addition to often being asymptomatic, TB detected through community-based screening may have its own spectrum of radiological features compared to TB diagnosed in clinics and hospitals, making it a distinct use-case for automated imaging algorithms. To date, CAD4TB has most frequently been used in healthcare centers to triage symptomatic patients^13,14,16,17,20,33,34^ with limited published data on its performance in symptom-agnostic screening programs. In this study, our decision to use an intentionally low CAD4TBv5 triaging threshold referred the majority (59.6%) of participants for sputum testing. Testing the sputum of participants with low CAD4TBv5 scores revealed that definite TB cases had a wide range of CXR abnormality (Figure 2 and 3). Over a third of the definite TB cases (33/99, 35.3%) had a CAD4TBv5 score lower than 60, which has been shown to be more than 80% sensitive for case finding in retrospective clinical settings.^17,24^ We considered whether the high HIV-prevalence in this study population may have lowered CAD4TB scores in the definite TB group. Previous studies have shown that CXRs of HIV-positive individuals with definite TB showed less TB-typical cavitation and more unspecific infiltrations.^22,23^ However we did not find evidence of different CAD4TB scores by HIV status, which may be due to the high rate of antiretroviral therapy use with resulting long-term viral suppression and immune reconstitution in our population.^35^ Another possible explanation for the wide range of CAD4TB scores among individuals with definite TB is that disease diagnosed through community-based screening represents an earlier “subclinical” stage of TB pathogenesis that is characterized by more subtle radiologic abnormalities on CXR. The existing CXR libraries^36^ that are available to train CAD algorithms use clinically-diagnosed TB as their references and thus may not be optimized to detect subclinical TB. CAD4TB is a proprietary algorithm and the demographic and clinical characteristics of the training data are unpublished.^37^ Therefore it is unclear to what extent community-diagnosed asymptomatic TB has been used to train CAD4TB. It would be interesting to see if CAD4TB and other CAD-algorithms can be refined to detect early, radiologically-subtle patterns of TB that is not yet clinically apparent. Increasing the transparency of algorithmic development could potentially speed up the refinement of CAD for this use.

A challenge to applying CAD for community-based screening is that it requires selection of a triaging threshold that is optimized for the study aims, number of available microbiological tests, and underlying TB prevalence.^13,20,25,33,34^ An inappropriately high triaging threshold choice may fail to capture all TB cases leading to underestimation of prevalence. A threshold that is too low may cause high costs and labor for laboratory diagnostics of sputum. In many cases it is necessary to conduct a pilot phase to optimize the triaging threshold for new settings. Ideally, this pilot should avoid selection of an empirical triage threshold and should instead test the sputum of all pilot participants to capture the full spectrum of active TB disease and provide unbiased data to determine an optimal triage threshold. Our experience with CAD4TBv5 and its successor CAD4TBv6 indicates that threshold adjustment is required when applying a new CAD4TB version. Threshold choice needs to be balanced between maximal case finding and cost-effectiveness. Due to cost constraints, public health settings are likely to favor a threshold that performs equivalently to an expert radiologist. We recommend that research programs consider a lower threshold that allows additional identification of radiologically subtle TB cases.

Limitations of our study include that only a single radiologist performed independent CXR reading which might have biased our analysis. As recommended by the WHO, the radiologist was instructed to overcall abnormality, but he may have been overly sensitive to very subtle abnormalities influencing the selection of our triaging threshold during the pilot stage. Further limitations are that only a single-spot sputum was the basis of microbiological data, which may have underestimated the number of definite TB cases. It is possible that we overestimated definite TB cases because of the inclusion of XpertUltra trace results. More research is necessary to determine whether individuals with XpertUltra “trace only” results represent early subclinical TB stages or if their results are false-positives. We attempted to address the uncertainty about “trace only” results using a sensitivity-analysis that excluded these cases (definite TB trace excluded) and did not find significantly different results. An additional limitation is that we did not collect sputum from participants who 1) were asymptomatic and had a CAD4TBv5 score below 25 (n=3,772), 2) were unable to produce sputum in the camp (n=1,112), and 3) could not be reached at home during a follow-up visit (n=54). The true TB status of those participants is unknown. The setting of our survey is a community with exceptionally high HIV-prevalence, which may limit the generalizability of our findings to other active case finding programs. A strength of our study is that the TB screening program was conducted as part of a multi-disease screening and bio-banking study within a demographic and health surveillance site.^38^ This means that the participants presented here have additional linked data describing clinical co-morbidities, banked bio-specimens from the time of the survey and are being actively followed through ongoing demographic and health surveillance activities.

During a multi-disease health screening in an HIV-endemic rural area we prospectively evaluated the ability of CAD4TB to interpret digital CXRs and triage participants for microbiological sputum testing. Piloting indicated that a low threshold would be required to achieve our goal of maximum active TB detection. We found that 1.0% of our population had microbiologically-confirmed TB and that these cases had a spectrum of radiological findings that ranged from highly abnormal to normal. Both CAD4TBv5 and CAD4TBv6 were able to achieve similar sensitivity to an expert radiologist in triaging CXRs for sputum collection, but only CAD4TBv6 also had a triaging threshold that achieved comparable specificity to the radiologist. Definite TB cases were largely asymptomatic (79.8%) and would have been missed in the absence of the digital CXR screening survey. With the caution that future applications may require setting-specific piloting to guide threshold selection, our data indicate that CAD4TB has the potential to replace human readers for CXR triaging in TB prevalence surveys.

## Methods

### Study design

The multi-disease community-based screening program ‘Vukuzazi’ used mobile vans to provide free health assessments in the rural uMkhanyakude district of KwaZulu-Natal in South Africa. The data for this analysis was collected during the first year of the project (between 25 May 2018 and 24 May 2019).^39^ Study field workers visited households to explain and provide the study description and invite eligible residents to participate. Eligibility criteria included a minimum age of 15 years and ongoing residency in the area. Participants provided written informed consent to participate in the study. In the case of participant age under 18, consent was also obtained from the parents or guardians. At the camp, participants answered questions in their preferred language about smoking, TB symptoms and history, HIV history and treatment, and the information was captured digitally.^38^ HIV status was assessed by 4th generation antibody/antigen test (Genscreen Ultra HIV Ag-Ab, Bio-Rad, Marnes-la-Coquette, France) on venous blood.

Posterior-anterior digital CXRs were obtained by a health worker using a mobile unit (Canon CXDI-NE) and the obtained digital CXRs were uploaded uncompressed in DICOM-format into a cloud storage. On a local workstation at the camp, the commercially-available software CAD4TBv5 calculated scores off-line for each CXR, indicating lung abnormalities and likelihood of active pulmonary tuberculosis^13,14,40^. CAD4TB’s methodology is based on initial lung field segmentation and subsequent analysis of the lung shape, symmetry, and costophrenic angles, resulting in an abnormality score between 0 and 100 (increasing with abnormality).^41^ The output score reflects the probability of active TB visible on the CXR.^14^ Following WHO guidelines for TB-prevalence surveys^10^, participants were referred for sputum examination if they reported any cardinal TB-symptom (fever, weight loss, cough, or night sweats) or if they had an abnormal CXR (indicated by a CAD4TBv5 score equals or higher than the triaging threshold in the camp). Sputum specimens were analyzed for Mtb using Xpert Ultra MTB/RIF® (XpertUltra) (Cepheid, Sunnyvale, CA, USA) and liquid BACTEC MGIT culture (MGIT) (Becton Dickinson, UK), held for 42 days. Within seven days of enrolment, an expert radiologist with more than 35 years of local experience reviewed all CXRs from high-resolution DICOM files on a computer screen in a central setting, blinded to CAD4TBv5 scores and any patient information. The radiologist categorized each CXR as 1) having either normal or abnormal lung fields and if abnormal 2) having radiological signs consistent with active TB or not.

### Obtaining a triaging threshold

Triaging with CAD4TB requires selecting a CAD4TB score as optimal triaging threshold. The aim of our survey was to maximize TB case-finding and to capture the full spectrum of active TB within the community. We conducted a pilot-phase with 1,090 participants who underwent chest radiography with an initially chosen CAD4TBv5 threshold of 60, seeking to adjust this threshold to match the radiologist’s ability of identifying abnormal lung fields. When planning the study, existing literature at that time showed CAD4TB thresholds around 60-65 to be more than 80% sensitive for active TB.^17,24^ Based on those we selected a CAD4TBv5 threshold of 60 to triage participants for sputum examination during the pilot phase, seeking to adjust this threshold after evaluating its performance. If during the pilot phase the expert radiologist indicated abnormal lung fields despite a CAD4TBv5 score below the threshold, a follow-up team contacted participants for sputum collection at home. After the pilot phase, we compared CAD4TBv5 scores to the radiologist’s assessment of abnormal lung fields and obtained threshold 25 which had a sensitivity of 85% to identify abnormal lung fields. CAD4TB threshold 25 was used to triage participants for the remainder of the study.

### Data analysis

CAD4TBv6, an updated version of the image interpretation software that uses deep neural networks,^14^ became available after data collection. CAD4TBv6 scores were calculated and analyzed retrospectively. We assessed the performance of CAD4TBv5 and v6 to identify any lung field abnormality like the radiologist. We compared the categorization of any lung field abnormality by the radiologist and CAD4TBv5 and v6 scores to diagnostic definitions of TB (definite TB, definite TB trace excluded and probable TB). Results are reported following Standards for Reporting of Diagnostic Accuracy (STARD).^42^ Performance was assessed by sensitivity, specificity, negative predictive value (NPV), positive predictive value (PPV), Area under the receiver operating curve (AUC) and area under precision-recall curve (PRAUC). Estimations of 95% confidence intervals (CI) are given. AUCs between CAD4TBv5 and v6 and HIV-positive and negative participants were compared using the DeLong method. For each CAD4TB threshold, we calculated the percentage (relative to all participants, n=9,914) and absolute number of participants who had an equal or higher score. Similar to Qin et al.,^19^ we calculated the number of needed sputum tests (NNT) which is required to find one person with definite TB and definite TB trace excluded and the number of missed TB cases for the radiologist and each CAD4TB score from 0-100. Missed TB cases were defined as the number of TB cases below each threshold. Missed TB cases were obtained for definite TB, definite TB trace excluded and probable TB definitions and stratified by symptom status. We compared CAD4TB scores between diagnostic groups of definite and probable TB, stratified by HIV status, and assessed significant differences using two-sided Mann-Whitney-Wilcoxon tests.

Data analysis was performed with R (version 4.0.3) using the packages ‘epiR’, ‘pROC’, and ‘precrec’.

## Data Availability

The Vukuzazi screening protocol and anonymized data can be requested at https://data.ahri.org/index.php/catalog/990.

## Data Availability

The Vukuzazi screening protocol as well as the dataset analyzed during the current study may be accessed via the AHRI Data Repository at https://data.ahri.org/index.php/catalog/990.^39^ upon approval of proposed analyses by the Vukuzazi Scientific Steering Committee and completion of a data access agreement.

## Code Availability statement

CAD4TB is a proprietary software and the code is not publically available. For questions about CAD4TB, please refer to Delft Imaging (https://www.delft.care/cad4tb/).

## Ethics declarations

Ethics approval was obtained from the University of KwaZulu-Natal Biomedical Research Ethics Committee (BE560/17), the London School of Hygiene & Tropical Medicine Ethics Committee (14722), and the Partners Institutional Review Board (2018P001802).

## Acknowledgments

‘Vukuzazi’ was the collective effort of a large team (see Supplementary Information). We thank the community of the uMkhanyakude district in KwaZulu-Natal for participating in this study. We acknowledge the members of the AHRI Community Advisory Board and the local and provincial Department of Health in their support of this project. We thank Delft for providing CAD4TBv6 scores free of charge. We dedicate this paper to the memories of Anand Ramnanan and Sibahle Gumbi who made a major contributions to the Vukuzazi Project.

The community-screening program ‘Vukuzazi’ is funded by the Africa Health Research Institute, the Wellcome Trust (201433/Z/16/Z), and the Bill and Melinda Gates Foundation (OPP1175182). Additional funders include NIAID (K08AI118538) and FIC (TW011687), National Institutes of Health, and Cascade IMPAc-TB Center (Contract # 75N93019C00070). The funders had no role in study design, data analysis and interpretation, or writing of the report. The mobile CXR unit including the real-time use of the CAD4TBv5 software was leased from Aurum Innova in partnership with Delft. The retrospective calculation of CAD4TBv6 scores was provided by Delft free of charge. Delft provided no funding and had no other role in this study. TN is supported through the South Africa Research Chairs Initiative (grant # 64809) and the Sub-Saharan African Network for TB/HIV Research Excellence (SANTHE), a DELTAS Africa Initiative [grant # DEL-15-006]. The DELTAS Africa Initiative is an independent funding scheme of the African Academy of Sciences (AAS)’s Alliance for Accelerating Excellence in Science in Africa (AESA) and supported by the New Partnership for Africa’s Development Planning and Coordinating Agency (NEPAD Agency) with funding from the Wellcome Trust [grant # 107752/Z/15/Z] and the UK government. The views expressed in this publication are those of the author(s) and not necessarily those of AAS, NEPAD Agency, Wellcome Trust or the UK government.

## Author Contributions

JF performed data analysis with figures and tables and writing of the report. SO and KB contributed to data analysis. DG contributed to data management. RG, AS, DP, OK, WH and MJS contributed to study design. TS SM, and TN contributed to the laboratory study setup and provision of test results. ADG contributed to study design and analysis conceptualization. SK and CL contributed to data analysis and supervision. EBW, study design, analysis conceptualization, revision and supervision. All authors contributed to the revision of the manuscript and approved the final version.

