## Supplemental Material for "Computer-aided interpretation of chest radiography reveals the spectrum of tuberculosis in rural South Africa"

### Supplemental Figures

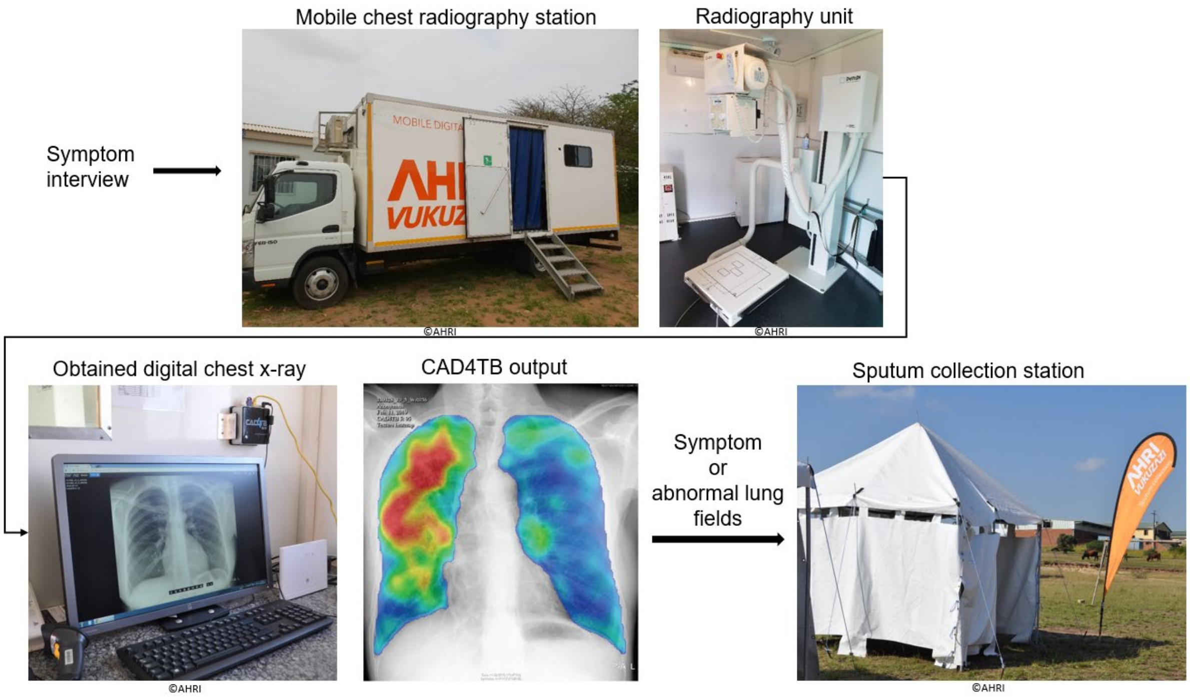

**Figure 1: Triaging for sputum assessment in Vukuzazi.** During the multi-morbidity health screening program Vukuzazi, 10,320 participants answered questions about current symptoms and underwent chest radiography in the mobile chest radiography station. The obtained digital chest x-rays were interpreted for present lung field abnormalities using the software CAD4TB. CAD4TB highlighted abnormal lung fields in the chest x-rays and presented those to the field radiographer. Participants were routed to sputum assessment station in the camp, if they reported TB-related symptoms (cough of any duration, fever, night sweat, or weight loss) or if CAD4TB detected lung field abnormalities. Photos ©AHRI.

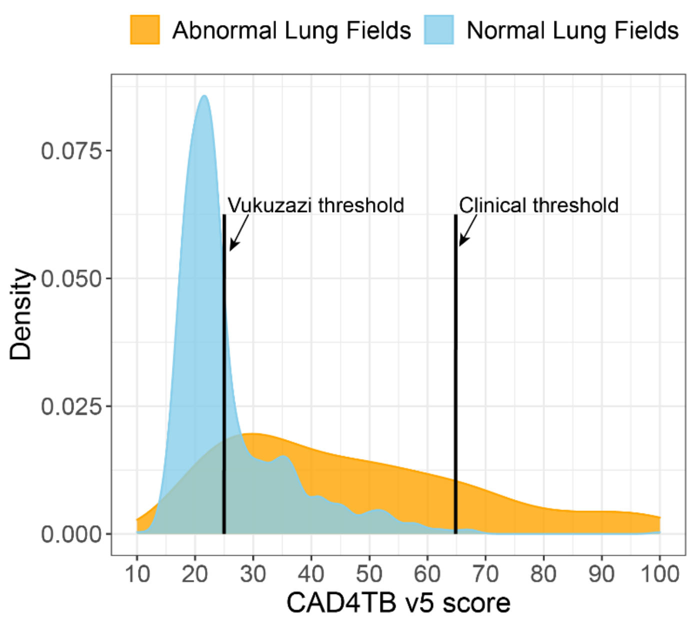


**Figure 2: Densities of CAD4TBv5 scores obtained during the pilot phase (n=1,090) stratified by the radiologist's classification of abnormal (n=198) and normal lung fields (n=892).** Triaging thresholds reported to be optimal in clinical thresholds are marked (65) next to the threshold chosen for this study (Vukuzazi) at 25.

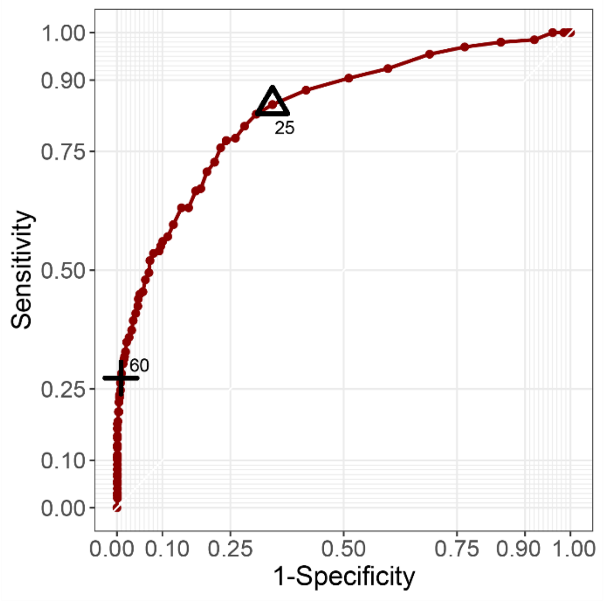


**Figure 3: Performance of CAD4TBv5 during the pilot-phase to detect CXRs classified as 'abnormal lung fields' by the radiologist.** CAD4TBv5 scores (n=1,090) were compared to the radiologist's classification of normal (n=892) and abnormal lung fields (n=198).

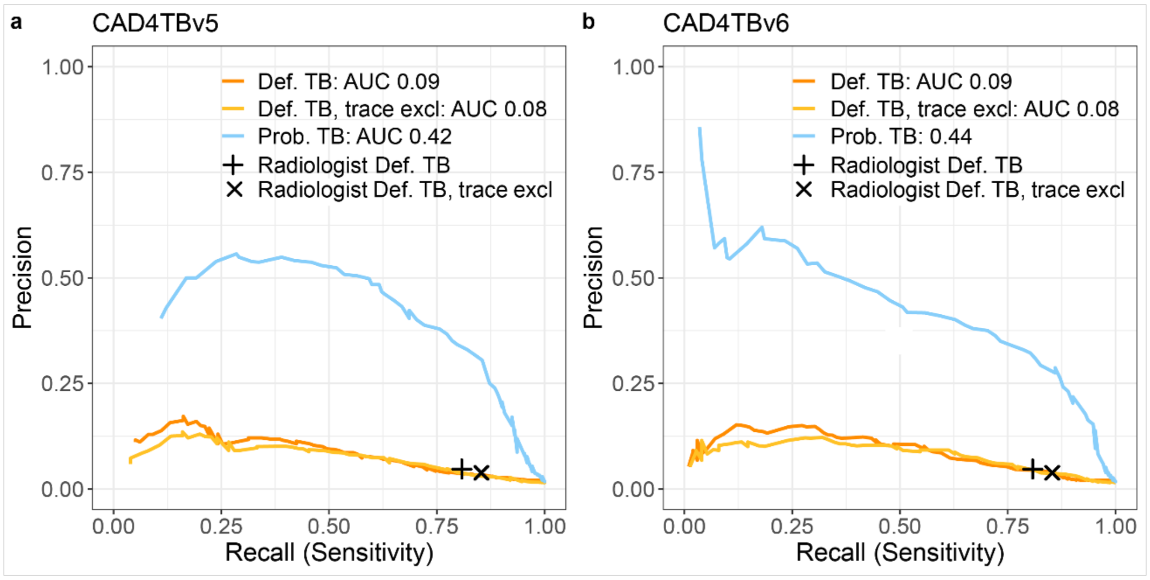


**Figure 4: Precision-Recall curve of CAD4TBv5 (a) and v6 (b) scores compared to diagnostic microbiological and/or radiological evidence.** Microbiological sputum test results were available for 4,976 participants. Radiological evidence was assessed among all participants who underwent chest radiography (n=9,914). Positive TB was defined as either definite TB with microbiological evidence definite TB excluding individuals that only had a XpertUltra trace result, or probable TB with radiological evidence of active TB but no microbiological evidence. The radiologist's precision and recall of detecting definite TB (+) and definite TB trace excluded is marked (x).

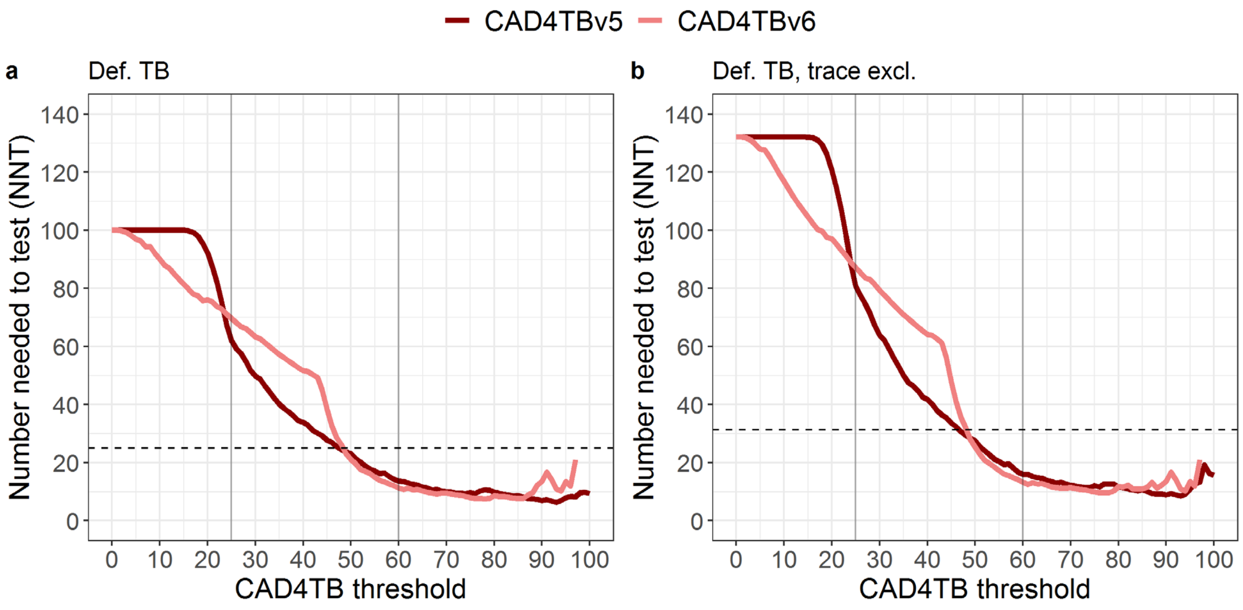


**Figure 5: Number needed to test (NNT) to find one participant with TB.** (a) The NNT to identify a participant with definite TB (n=99) or (b) definite TB trace excluded (n=75). NNT is the number of participants with a CAD4TB score equal or above each threshold divided by the number of identified definite TB at the respective threshold. The dashed line indicates the NNT of the radiologist (a: 25, b: 31).

**Supplemental Tables**

**Table 1: Performance of the radiologist and CAD4TBv5 and v6 to identify definite TB.** Performance among participants with microbiological sputum test results (n=4,976). Definite TB was defines as either positive XpertUltra or liquid culture test result (n=99). Performance is given as sensitivity, specificity, positive predictive value (PPV) and negative predictive value (NPV) in % with 95% confidence intervals (CI), number of participants need to test and missed definite TB cases. Further listed are the percentage of participants who required sputum testing due to lung field abnormality, deemed by the radiologist and CAD4TB and the number of needed tests (NNT) to find one participant with definite TB. Numbers of missed definite TB cases (all and asymptomatic) are listed as absolute numbers and relative to all definite TB cases (n=99).

| | **Sensitivity % (CI)** | | **Specificity % (CI)** | | **PPV % (CI)** | | **NPV % (CI)** | | **Participants requiring sputum testing (%)** | | **NNT** | | **#missed definite TB (%)** | | **#missed asymptomatic definite TB (%)** | |
|---|---|---|---|---|---|---|---|---|---|---|---|---|---|---|---|---|
| **Radiologist:** Any CXR abnormality | 80.8 (71.7-88.0) | | 66.9 (65.6-68.2) | | 4.7 (3.8-5.8) | | 99.4 (99.1-99.7) | | 2,002 (20.2) | | 25 | | 19 (19.2) | | 15 (15.2) | |
| CXR diag. active TB | 30.3 (21.5-40.4) | | 97.0 (96.5-97.4) | | 16.9 (11.7-23.3) | | 98.6 (98.2-98.9) | | 202 (2.0) | | 6.7 | | 69 (69.7) | | 55 (55.6) | |
| **CAD4TB** | **v5** | **v6** | **v5** | **v6** | **v5** | **v6** | **v5** | **v6** | **v5** | **v6** | **v5** | **v6** | **v5** | **v6** | **v5** | **v6** |
| 20 | 99.0 (94.5-100) | 92.9 (86-97.1) | 1.4 (1.1-1.7) | 11.7 (10.8-12.6) | 2.0 (1.6-2.4) | 2.1 (1.7-2.6) | 98.5 (92.1-100) | 98.8 (97.5-99.5) | 9,047 (91.3) | 6,990 (70.5) | 92 | 76 | 1 (1.0) | 7 (7.0) | 0 (0.0) | 5 (5.1) |
| 25 | 96.0 (90.0-98.9) | 90.9 (83.4-95.8) | 8.0 (7.3-8.8) | 16.9 (15.9-18) | 2.1 (1.7-2.5) | 2.2 (1.8-2.7) | 99 (97.4-99.7) | 98.9 (98-99.5) | 5,906 (59.6) | 6,276 (63.3) | 62 | 70 | 4 (4.0) | 9 (9.1) | 1 (1.0) | 6 (6.1) |
| 30 | 91.9 (84.7-96.4) | 89.9 (82.2-95) | 28.7 (27.5-30.0) | 22.4 (21.2-23.6) | 2.6 (2.1-3.1) | 2.3 (1.8-2.8) | 99.4 (98.9-99.8) | 99.1 (98.3-99.6) | 4,532 (45.7) | 5,635 (56.84) | 50 | 63 | 8 (8.1) | 10 (10.1) | 5 (5.1) | 7 (7.1) |

| | | | | | | | | | | | | | | | |
|---|---|---|---|---|---|---|---|---|---|---|---|---|---|---|---|
| 35 | 86.9 (78.6-92.8) | 88.9 (81-94.3) | 45.6 (44.2-47) | 28 (26.7-29.3) | 3.1 (2.5-3.9) | 2.4 (2.0-3.0) | 99.4 (99-99.7) | 99.2 (98.6-99.6) | 3,459 (34.8) | 5,042 (50.9) | 40 | 57 | 13 (13.1) | 11 (11.1) | 10 (10.1) | 8 (8.1) |
| 40 | 79.8 (70.5-87.2) | 88.9 (81.0-94.3) | 57.4 (56-58.8) | 33.3 (31.9-34.6) | 3.7 (2.9-4.5) | 2.6 (2.1-3.2) | 99.3 (98.9-99.6) | 99.3 (98.8-99.7) | 2,677 (27.0) | 4,550 (45.9) | 34 | 52 | 20 (20.2) | 11 (11.1) | 14 (14.1) | 8 (8.1) |
| 41 | 77.8 (68.3-85.5) | 87.9 (79.8-93.6) | 59.7 (58.4-61.1) | 34.2 (32.8-35.5) | 3.8 (3.0-4.7) | 2.6 (2.1-3.2) | 99.3 (98.9-99.5) | 99.3 (98.8-99.6) | 2,533 (25.5) | 4,468 (45.1) | 33 | 51 | 22 (22.2) | 12 (12.1) | 16 (16.2) | 9 (9.1) |
| 42 | 77.8 (68.3-85.5) | 87.9 (79.8-93.6) | 61.7 (60.3-63.1) | 35.1 (33.7-36.4) | 4.0 (3.1-4.9) | 2.7 (2.1-3.3) | 99.3 (98.9-99.5) | 99.3 (98.8-99.6) | 2,397 (24.2) | 4,387 (44.3) | 31 | 50 | 22 (22.2) | 12 (12.1) | 16 (16.2) | 9 (9.1) |
| 43 | 75.8 (66.1-83.8) | 87.9 (79.8-93.6) | 63.6 (62.3-65) | 36.2 (34.8-37.5) | 4.1 (3.2-5.1) | 2.7 (2.1-3.3) | 99.2 (98.9-99.5) | 99.3 (98.8-99.6) | 2,264 (22.8) | 4,288 (43.3) | 30 | 49 | 24 (24.2) | 12 (12.1) | 17 (17.2) | 9 (9.1) |
| 44 | 74.7 (65.0-82.9) | 87.9 (79.8-93.6) | 65.3 (64-66.7) | 40.6 (39.2-42) | 4.2 (3.3-5.2) | 2.9 (2.3-3.6) | 99.2 (98.9-99.5) | 99.4 (99-99.7) | 2,167 (21.9) | 3,927 (39.6) | 29 | 45 | 25 (25.3) | 12 (12.1) | 18 (18.2) | 9 (9.1) |
| 45 | 74.7 (65.0-82.9) | 86.9 (78.6-92.8) | 67.2 (65.9-68.5) | 48.8 (47.4-50.3) | 4.4 (3.5-5.5) | 3.3 (2.7-4.1) | 99.2 (98.9-99.5) | 99.5 (99.1-99.7) | 2,051 (20.7) | 3,295 (33.2) | 28 | 38 | 25 (25.3) | 13 (13.1) | 18 (18.2) | 10 (10.1) |
| 46 | 72.7 (62.9-81.2) | 85.9 (77.4-92) | 68.9 (67.6-70.2) | 55.8 (54.4-57.2) | 4.5 (3.6-5.7) | 3.8 (3.0-4.7) | 99.2 (98.8-99.5) | 99.5 (99.1-99.7) | 1,943 (19.6) | 2,799 (28.2) | 27 | 33 | 27 (27.3) | 14 (14.1) | 20 (20.2) | 11 (11.1) |
| 47 | 72.7 (62.9-81.2) | 82.8 (73.9-89.7) | 70.5 (69.2-71.8) | 62.6 (61.2-64) | 4.8 (3.8-6.0) | 4.3 (3.4-5.3) | 99.2 (98.8-99.5) | 99.4 (99.1-99.7) | 1,842 (18.6) | 2,348 (23.7) | 26 | 29 | 27 (27.3) | 17 (17.2) | 20 (20.2) | 13 (13.1) |

| | | | | | | | | | | | | | | | | |
|---|---|---|---|---|---|---|---|---|---|---|---|---|---|---|---|---|
| 48 | 72.7 (62.9-81.2) | 76.8 (67.2-84.7) | 71.8 (70.5-73.1) | 68.0 (66.7-69.3) | 5.0 (3.9-6.2) | 4.6 (3.7-5.8) | 99.2 (98.8-99.5) | 99.3 (99-99.6) | 1,762 (17.8) | 1,997 (20.1) | 25 | 26 | 27 (27.3) | 23 (23.2) | 20 (20.2) | 18 (18.2) |
| 49 | 71.7 (61.8-80.3) | 74.7 (65-82.9) | 73.2 (72-74.5) | 72.6 (71.4-73.9) | 5.2 (4-6.5) | 5.3 (4.1-6.6) | 99.2 (98.8-99.5) | 99.3 (99-99.6) | 1,684 (17.0) | 1,701 (17.2) | 24 | 23 | 28 (28.3) | 25 (25.3) | 21 (21.2) | 19 (19.2) |
| 50 | 69.7 (59.6-78.5) | 70.7 (60.7-79.4) | 74.5 (73.3-75.8) | 76.4 (75.1-77.5) | 5.3 (4.1-6.6) | 5.7 (4.5-7.2) | 99.2 (98.8-99.4) | 99.2 (98.9-99.5) | 1,598 (16.1) | 1,474 (14.9) | 23 | 21 | 30 (30.3) | 29 (29.3) | 23 (23.2) | 22 (22.2) |
| 60 | 55.6 (45.2-65.5) | 55.6 (45.2-65.5) | 87.9 (87.0-88.8) | 89.9 (89.0-90.7) | 8.6 (6.5-11) | 10.1 (7.7-12.9) | 99.0 (98.6-99.3) | 99.0 (98.7-99.3) | 751 (7.6) | 623 (6.3) | 14 | 11 | 44 (44.4) | 44 (44.4) | 35 (35.4) | 34 (34.3) |
| 70 | 40.4 (30.7-50.7) | 36.4 (26.9-46.6) | 93.5 (92.8-94.2) | 94.6 (93.9-95.2) | 11.3 (8.2-15.0) | 12.0 (8.5-16.2) | 98.7 (98.4-99.0) | 98.7 (98.3-99.0) | 401 (4.0) | 342 (3.4) | 10 | 10 | 59 (59.6) | 63 (63.6) | 45 (45.5) | 49 (49.5) |
| 80 | 24.2 (16.2-33.9) | 19.2 (12.0-28.3) | 96.2 (95.6-96.7) | 97.4 (97.0-97.9) | 11.5 (7.5-16.6) | 13.2 (8.1-19.8) | 98.4 (98.0-98.8) | 98.3 (97.9-98.7) | 234 (2.4) | 165 (1.7) | 10 | 9 | 75 (75.8) | 80 (80.8) | 60 (60.6) | 64 (64.6) |
| 90 | 19.2 (12.0-28.3) | 4.0 (1.1-10) | 97.9 (97.5-98.3) | 99.1 (98.8-99.3) | 16.0 (9.9-23.8) | 8.3 (2.3-20.0) | 98.4 (98.0-98.7) | 98.1 (97.6-98.4) | 133 (1.3) | 54 (0.5) | 7 | 14 | 80 (80.8) | 95 (96.0) | 65 (65.7) | 75 (75.8) |

**Table 2: Performance of the radiologist and CAD4TBv5 and v6 to identify definite TB trace excluded.** Performance among participants with microbiological sputum test results, excluding participants who were only definite TB positive due to a XpertUltra trace result (remaining n=4,952). Performance is given as sensitivity, specificity, positive predictive value (PPV) and negative predictive value (NPV) in % with 95% confidence intervals (CI). Further listed are the percentage of participants who required sputum testing due to lung field abnormality, deemed by the radiologist and CAD4TB and the number of needed tests (NNT) to find one participant with definite TB, trace excluded. Numbers of missed definite TB cases (all and asymptomatic) are listed as absolute numbers and relative to all definite TB cases trace excluded (n=75).

| | **Sensitivity % (CI)** | | **Specificity % (CI)** | | **PPV % (CI)** | | **NPV % (CI)** | | **Participants requiring sputum testing (%)** | | **NNT** | | **#missed definite TB, trace excluded (%)** | | **#missed asymptomatic, trace excluded (%)** | |
|---|---|---|---|---|---|---|---|---|---|---|---|---|---|---|---|---|
| **Radiologist**: Any CXR abnormality | 85.3 (75.3-92.4) | | 66.9 (65.6-68.2) | | 3.8 (3.0-4.8) | | 99.7 (99.4-99.8) | | 2,002 (20.2) | | 31 | | 11 (14.7) | | 9 (12.0) | |
| CXR diag. active TB | 33.8 (23.0-46.0) | | 97.0 (96.5-97.4) | | 14.0 (9.2-20.2) | | 99.0 (98.7-99.3) | | 202 (2.0) | | 8 | | 50 (50.5) | | 41 (41.4) | |
| **CAD4TB score** | **v5** | **v6** | **v5** | **v6** | **v5** | **v6** | **v5** | **v6** | **v5** | **v6** | **v5** | **v6** | **v5** | **v6** | **v5** | **v6** |
| 20 | 100 (95.2-100) | 96 (88.8-99.2) | 1.4 (1.1-1.7) | 11.7 (10.8-12.6) | 1.5 (1.2-1.9) | 1.6 (1.3-2.1) | 100 (94.6-100) | 99.5 (98.5-99.9) | 9,047 (91.3) | 6,990 (70.5) | 121 | 97 | 0 (0.0) | 3 (4.0) | 0 (0.0) | 2 (2.7) |
| 25 | 97.3 (90.7-99.7) | 96.0 (88.8-99.2) | 8.0 (7.3-8.8) | 16.9 (15.9-18) | 1.6 (1.3-2) | 1.7 (1.4-2.2) | 99.5 (98.2-99.9) | 99.6 (99.1-99.9) | 5,906 (59.6) | 6,276 (63.3) | 81 | 87 | 2 (2.7) | 3 (4.0) | 1 (1.3) | 2 (2.7) |
| 30 | 94.7 (86.9-98.5) | 94.7 (86.9-98.5) | 28.7 (27.5-30.0) | 22.4 (21.2-23.6) | 2.0 (1.6-2.5) | 1.8 (1.4-2.3) | 99.7 (99.3-99.9) | 99.6 (99.1-99.9) | 4,532 (45.7) | 5,635 (56.8) | 64 | 79 | 4 (5.3) | 4 (5.3) | 3 (4.0) | 3 (4.0) |
| 35 | 92.0 (83.4-97.0) | 94.7 (86.9-98.5) | 45.6 (44.2-47.0) | 28.0 (26.7-29.3) | 2.5 (2-3.2) | 2.0 (1.6-2.5) | 99.7 (99.4-99.9) | 99.7 (99.2-99.9) | 3,459 (34.8) | 5,042 (50.9) | 50 | 71 | 6 (8.0) | 4 (5.3) | 5 (6.7) | 3 (4.0) |
| 40 | 85.3 (75.3-92.4) | 94.7 (86.9-98.5) | 57.4 (56-58.8) | 33.3 (31.9-34.6) | 3.0 (2.3-3.8) | 2.1 (1.7-2.7) | 99.6 (99.3-99.8) | 99.8 (99.4-99.9) | 2,677 (27.0) | 4,550 (45.9) | 42 | 64 | 11 (14.7) | 4 (5.3) | 8 (10.7) | 3 (4.0) |
| 45 | 81.3 (70.7-89.4) | 92.0 (83.4-97.0) | 67.2 (65.9-68.5) | 48.8 (47.4-50.3) | 3.7 (2.8-4.7) | 2.7 (2.1-3.4) | 99.6 (99.3-99.8) | 99.7 (99.5-99.9) | 2,051 (20.7) | 3,295 (33.2) | 34 | 48 | 14 (18.7) | 6 (8.0) | 11 (14.7) | 5 (6.7) |
| 50 | 77.3 (66.2-86.2) | 77.3 (66.2-86.2) | 74.5 (73.3-75.8) | 76.4 (75.1-77.5) | 4.5 (3.4-5.7) | 4.8 (3.7-6.1) | 99.5 (99.3-99.7) | 99.5 (99.3-99.7) | 1,598 (16.1) | 1,474 (14.9) | 23 | 21 | 17 (22.7) | 17 (22.7) | 14 (18.7) | 13 (17.3) |

| | | | | | | | | | | | | | | | | |
|---|---|---|---|---|---|---|---|---|---|---|---|---|---|---|---|---|
| 60 | 62.7 (50.7-73.6) | 62.7 (50.7-73.6) | 87.9 (87-88.8) | 89.9 (89-90.7) | 7.4 (5.5-9.7) | 8.7 (6.5-11.4) | 99.4 (99.1-99.6) | 99.4 (99.1-99.6) | 751 (7.6) | 623 (6.3) | 14 | 11 | 28 (37.3) | 28 (37.3) | 23 (30.7) | 22 (29.3) |
| 70 | 44.0 (32.5-55.9) | 40.0 (28.9-52.0) | 93.5 (92.8-94.2) | 94.6 (93.9-95.2) | 9.5 (6.6-13.1) | 10.2 (7.0-14.2) | 99.1 (98.8-99.3) | 99.0 (98.7-99.3) | 401 (4.0) | 342 (3.4) | 10 | 10 | 42 (56.0) | 45 (60.0) | 32 (42.7) | 35 (46.7) |
| 80 | 26.7 (17.1-38.1) | 18.7 (10.6-29.3) | 96.2 (95.6-96.7) | 97.4 (97-97.9) | 9.8 (6.1-14.7) | 10.1 (5.6-16.3) | 98.8 (98.5-99.1) | 98.7 (98.4-99) | 234 (2.4) | 165 (1.7) | 10 | 9 | 55 (73.3) | 61 (81.3) | 44 (58.7) | 49 (65.3) |
| 90 | 20.0 (11.6-30.8) | 5.3 (1.5-13.1) | 97.9 (97.5-98.3) | 99.1 (98.8-99.3) | 13.0 (7.5-20.6) | 8.3 (2.3-20.0) | 98.8 (98.4-99.1) | 98.6 (98.2-98.9) | 133 (1.3) | 54 (0.5) | 7 | 14 | 60 (80.0) | 71 (94.7) | 49 (65.3) | 56 (74.7) |

**Table S3: Performance of CAD4TBv5 to identify probable TB.** Performance among participants who underwent chest radiography (n=9,914). Probable TB was defined as radiological evidence indicated by the radiologist by 'CXR diagnostic of active TB' but no microbiological evidence (n=172). Performance is given as sensitivity, specificity, positive predictive value (PPV) and negative predictive value (NPV) in % with 95% confidence intervals (CI). The numbers of missed probable TB cases (all and asymptomatic) are listed as absolute numbers absolute and relative to all probable TB cases (n=172).

| **CAD4TB score** | **Sensitivity % (CI)** | | **Specificity % (CI)** | | **PPV % (CI)** | | **NPV % (CI)** | | **#missed probable TB (%)** | | **#missed asymptomatic probable TB (%)** | |
|---|---|---|---|---|---|---|---|---|---|---|---|---|
| | **v5** | **v6** | **v5** | **v6** | **v5** | **v6** | **v5** | **v6** | **v5** | **v6** | **v5** | **v6** |
| 20 | 100 (97.9-100) | 99.4 (96.8-100) | 8.9 (8.3-9.5) | 30.0 (29.1-30.9) | 1.9 (1.6-2.2) | 2.4 (2.1-2.8) | 100 (99.6-100) | 100 (99.8-100) | 0 (0.0) | 1 (0.6) | 0 (0.0) | 1 (0.6) |
| 25 | 99.4 (96.8-100) | 98.8 (95.9-99.9) | 41.1 (40.2-42.1) | 37.3 (36.4-38.3) | 2.9 (2.5-3.4) | 2.7 (2.3-3.1) | 100 (99.9-100) | 99.9 (99.8-100) | 1 (0.6) | 2 (1.2) | 1 (0.6) | 2 (1.2) |
| 30 | 97.7 (94.2-99.4) | 98.8 (95.9-99.9) | 55.2 (54.2-56.2) | 43.9 (42.9-44.9) | 3.7 (3.2-4.3) | 3 (2.6-3.5) | 99.9 (99.8-100) | 100 (99.8-100) | 4 (2.3) | 2 (1.2) | 4 (2.3) | 2 (1.2) |
| 35 | 97.1 (93.3-99) | 98.3 (95.0-99.6) | 66.2 (65.3-67.1) | 50.0 (49.0-51.0) | 4.8 (4.1-5.6) | 3.4 (2.9-3.9) | 99.9 (99.8-100) | 99.9 (99.8-100) | 5 (2.9) | 3 (1.7) | 5 (2.9) | 3 (1.7) |
| 40 | 95.9 (91.8-98.3) | 98.3 (95.0-99.6) | 74.2 (73.3-75.1) | 55.0 (54.0-56.0) | 6.2 (5.3-7.1) | 3.7 (3.2-4.3) | 99.9 (99.8-100) | 99.9 (99.8-100) | 7 (4.1) | 3 (1.7) | 6 (3.5) | 3 (1.7) |
| 45 | 95.3 (91.0-98.0) | 97.7 (94.2-99.4) | 80.6 (79.8-81.4) | 67.9 (67-68.8) | 8.0 (6.9-9.3) | 5.1 (4.4-5.9) | 99.9 (99.8-100) | 99.9 (99.8-100) | 8 (4.7) | 4 (1.7) | 7 (4.1) | 4 (2.3) |
| 50 | 93.6 (88.8-96.8) | 95.3 (91.0-98.0) | 85.2 (84.5-85.9) | 86.6 (85.9-87.2) | 10.1 (8.6-11.7) | 11.1 (9.6-12.8) | 99.9 (99.8-99.9) | 99.9 (99.8-100) | 11 (6.4) | 8 (4.7) | 10 (5.8) | 7 (4.1) |
| 60 | 90.1 (84.6-94.1) | 87.8 (81.9-92.3) | 93.9 (93.4-94.4) | 95.2 (94.7-95.6) | 20.6 (17.8-23.7) | 24.2 (20.9-27.8) | 99.8 (99.7-99.9) | 99.8 (99.7-99.9) | 17 (9.9) | 21 (12.2) | 14 (8.1) | 18 (10.5) |
| 70 | 79.7 (72.9-85.4) | 72.1 (64.8-78.7) | 97.3 (96.9-97.6) | 97.8 (97.4-98.0) | 34.2 (29.5-39) | 36.3 (31.2-41.6) | 99.6 (99.5-99.7) | 99.5 (99.3-99.6) | 35 (20.3) | 48 (27.9) | 27 (15.7) | 39 (22.7) |

| | | | | | | | | | | | | |
|---|---|---|---|---|---|---|---|---|---|---|---|---|
| 80 | 62.8 (55.1-70) | 44.8 (37.2-52.5) | 98.7 (98.5-98.9) | 99.1 (98.9-99.3) | 46.2 (39.6-52.8) | 46.7 (38.9-54.6) | 99.3 (99.2-99.5) | 99.0 (98.8-99.2) | 64 (37.2) | 95 (55.2) | 46 (26.7) | 68 (39.5) |
| 90 | 41.9 (34.4-49.6) | 18.6 (13.1-25.2) | 99.4 (99.2-99.5) | 99.8 (99.7-99.9) | 54.1 (45.3-62.8) | 59.3 (45-72.4) | 99.0 (98.8-99.2) | 98.6 (98.3-98.8) | 100 (58.1) | 140 (81.4) | 74 (43.0) | 104 (60.5) |

**Table 3: Performance of CAD4TBv5 to identify probable TB.** Probable TB was defined as radiological evidence indicated by the radiologist by 'CXR diagnostic of active TB' but no microbiological evidence. Performance is given as sensitivity, specificity, positive predictive value (PPV) and negative predictive value (NPV) in % with 95% confidence intervals (CI). The number of needed tests (NNT) is reported in absolute numbers and relative to all (n=9,914) participants. The number of all and asymptomatic missed definite TB cases is listed in absolute numbers and relative to all probable TB cases (n=172).

| **CAD4TB score** | **Sensitivity % (CI)** | | **Specificity % (CI)** | | **PPV % (CI)** | | **NPV % (CI)** | | **#missed probable TB (%)** | | **#missed asymptomatic probable TB (%)** | |
|---|---|---|---|---|---|---|---|---|---|---|---|---|
| | **v5** | **v6** | **v5** | **v6** | **v5** | **v6** | **v5** | **v6** | **v5** | **v6** | **v5** | **v6** |
| 20 | 100 (97.9-100) | 99.4 (96.8-100) | 8.9 (8.3-9.5) | 30.0 (29.1-30.9) | 1.9 (1.6-2.2) | 2.4 (2.1-2.8) | 100 (99.6-100) | 100 (99.8-100) | 0 (0.0) | 1 (0.6) | 0 (0.0) | 1 (0.6) |
| 25 | 99.4 (96.8-100) | 98.8 (95.9-99.9) | 41.1 (40.2-42.1) | 37.3 (36.4-38.3) | 2.9 (2.5-3.4) | 2.7 (2.3-3.1) | 100 (99.9-100) | 99.9 (99.8-100) | 1 (0.6) | 2 (1.2) | 1 (0.6) | 2 (1.2) |
| 30 | 97.7 (94.2-99.4) | 98.8 (95.9-99.9) | 55.2 (54.2-56.2) | 43.9 (42.9-44.9) | 3.7 (3.2-4.3) | 3 (2.6-3.5) | 99.9 (99.8-100) | 100 (99.8-100) | 4 (2.3) | 2 (1.2) | 4 (2.3) | 2 (1.2) |
| 35 | 97.1 (93.3-99) | 98.3 (95.0-99.6) | 66.2 (65.3-67.1) | 50.0 (49.0-51.0) | 4.8 (4.1-5.6) | 3.4 (2.9-3.9) | 99.9 (99.8-100) | 99.9 (99.8-100) | 5 (2.9) | 3 (1.7) | 5 (2.9) | 3 (1.7) |
| 40 | 95.9 (91.8-98.3) | 98.3 (95.0-99.6) | 74.2 (73.3-75.1) | 55.0 (54.0-56.0) | 6.2 (5.3-7.1) | 3.7 (3.2-4.3) | 99.9 (99.8-100) | 99.9 (99.8-100) | 7 (4.1) | 3 (1.7) | 6 (3.5) | 3 (1.7) |
| 45 | 95.3 (91.0-98.0) | 97.7 (94.2-99.4) | 80.6 (79.8-81.4) | 67.9 (67-68.8) | 8.0 (6.9-9.3) | 5.1 (4.4-5.9) | 99.9 (99.8-100) | 99.9 (99.8-100) | 8 (4.7) | 4 (1.7) | 7 (4.1) | 4 (2.3) |
| 50 | 93.6 (88.8-96.8) | 95.3 (91.0-98.0) | 85.2 (84.5-85.9) | 86.6 (85.9-87.2) | 10.1 (8.6-11.7) | 11.1 (9.6-12.8) | 99.9 (99.8-99.9) | 99.9 (99.8-100) | 11 (6.4) | 8 (4.7) | 10 (5.8) | 7 (4.1) |
| 60 | 90.1 (84.6-94.1) | 87.8 (81.9-92.3) | 93.9 (93.4-94.4) | 95.2 (94.7-95.6) | 20.6 (17.8-23.7) | 24.2 (20.9-27.8) | 99.8 (99.7-99.9) | 99.8 (99.7-99.9) | 17 (9.9) | 21 (12.2) | 14 (8.1) | 18 (10.5) |
| 70 | 79.7 (72.9-85.4) | 72.1 (64.8-78.7) | 97.3 (96.9-97.6) | 97.8 (97.4-98.0) | 34.2 (29.5-39) | 36.3 (31.2-41.6) | 99.6 (99.5-99.7) | 99.5 (99.3-99.6) | 35 (20.3) | 48 (27.9) | 27 (15.7) | 39 (22.7) |

| | | | | | | | | | | | | |
|---|---|---|---|---|---|---|---|---|---|---|---|---|
| 80 | 62.8 (55.1-70) | 44.8 (37.2-52.5) | 98.7 (98.5-98.9) | 99.1 (98.9-99.3) | 46.2 (39.6-52.8) | 46.7 (38.9-54.6) | 99.3 (99.2-99.5) | 99.0 (98.8-99.2) | 64 (37.2) | 95 (55.2) | 46 (26.7) | 68 (39.5) |
| 90 | 41.9 (34.4-49.6) | 18.6 (13.1-25.2) | 99.4 (99.2-99.5) | 99.8 (99.7-99.9) | 54.1 (45.3-62.8) | 59.3 (45-72.4) | 99.0 (98.8-99.2) | 98.6 (98.3-98.8) | 100 (58.1) | 140 (81.4) | 74 (43.0) | 104 (60.5) |

**Table 4: Area under the receiver operating curves (AUC) from CAD4TBv5 and v6 compared to diagnostic microbiological and/or radiological evidence.** Positive TB was defined as either definite TB with microbiological evidence, definite TB excluding samples that only had a XpertUltra trace result, or probable TB with radiological signs of TB but no microbiological evidence.

| | **Definite TB** | **Definite TB trace excl.** | **Probable TB** |
|---|---|---|---|
| v5 AUC (CI) | 0.78 (0.73-0.83) | 0.82 (0.77-0.87) | 0.96 (0.95-0.98) |
| v5 AUC (CI) HIV-negative | 0.75 (0.68-0.83) | 0.81 (0.74-0.89) | 0.96 (0.93-0.98) |
| v5 AUC (CI) HIV-positive | 0.80 (0.72-0.87) | 0.82 (0.74-0.89) | 0.97 (0.96-0.99) |
| v6 AUC (CI) | 0.79 (0.73-0.84) | 0.84 (0.79-0.89) | 0.96 (0.95-0.98) |
| v6 AUC (CI) HIV-negative | 0.76 (0.68-0.84) | 0.83 (0.76-0.91) | 0.95 (0.93-0.98) |
| v6 AUC (CI) HIV-positive | 0.81 (0.74-0.88) | 0.82 (0.76-0.89) | 0.97 (0.97-0.98) |

**Table 5: P-values of comparing the diagnostic performance of CAD4TB to gold standards measured in area under the curve (AUC) between diagnostic groups.**

| | CAD4TBv5 | CAD4TBv6 |
|---|---|---|
| Definite TB vs. Definite TB, trace excluded | 0.28 | 0.18 |
| Definite TB vs. Probable TB | $5.25x10^{-11}$ | $9.05x10^{-10}$ |
| Definite TB, trace excluded vs. Probable TB | $4.51x10^{-7}$ | $2.5x10^{-6}$ |

**Table 6: P-values of comparing the diagnostic performance of CAD4TB to gold standards measured in area under the curve (AUC) between HIV-positive and negative individuals.** Gold standards were defined as either definite TB with microbiological evidence (M+), definite TB excluding M+ samples that only had a XpertUltra trace result, or probable TB with no microbiological evidence but radiological signs of TB (M-/0 R+). CAD4TB scores from version 5 (v5) and 6 (v6) were compared against these gold standards.

| **Gold standard** | **CAD4TBv5** | **CAD4TBv6** |
|---|---|---|
| Def. TB | 0.42 | 0.32 |
| Def. TB trace excl. | 0.96 | 0.85 |
| Prob. TB | 0.30 | 0.16 |

***Vukuzazi Team: Staff who significantly contributed to the implementation and conduct of Vukuzazi.**

* Denotes team members who were closely involved with the design, implementation and oversight of Vukuzazi.

| **Name** | **Role** |
|---|---|
| *Alison Grant[2,4,6,10] | Co-investigator |
| Anand Ramnanan[2] | Procurement |
| Anele Mkhwanazi[2] | Clinical Research Assistant Supervisor |
| Antony Rapulana[2] | Laboratory Technologist |
| Anupa Singh[2] | Laboratory Technician |
| Ashentha Govender[2] | Laboratory Technician |
| *Ashmika Surujdeen[2] | Study Coordinator |
| Ayanda Zungu[2] | Clinical Research Assistant |
| Boitsholo Mfolo[14] | Radiographer |
| Bongani Magwaza[2] | General Worker |
| Bongumenzi Ndlovu[2] | Enrolled Nurse |
| Clive Mavimbela[2] | Operational Oversight |
| Costa Criticos[2] | Operational Oversight |
| *Day Munatsi[2] | Head: Research Data Systems |
| *Deenan Pillay[2,5] | Principal Investigator (2017-2019) |
| *Dickman Gareta[2] | Head: Research Data Management |
| Dilip Kalyan[2] | Operational Oversight |
| Doctar Mlambo[2] | Enrolled Nurse |
| *Emily Wong[2,11,12,13] | Co-Principal Investigator |
| Fezeka Mfeka[2] | Clinical Research Assistant |
| Freddy Mabetlela[2] | Laboratory Technologist |
| *Gregory Ording-Jespersen[2] | Laboratory Data Supervisor |
| Hannah Keal[2] | Communications |
| Hlengiwe Dlamini[2] | Enrolled Nurse |

| | |
|---|---|
| Hlengiwe Khathi[2] | Biorepository Research Assistant |
| Hlobisile Chonco[2] | Enrolled Nurse |
| Hlobisile Gumede[2] | Clinical Research Assistant |
| Hlobisile Gumede[2] | Clinical Research Assistant |
| Hlolisile Khumalo[2] | Nursing Manager |
| Hloniphile Ngubane[2] | Professional Nurse |
| *Hollis Shen[2] | Head: Exploratory Research Division |
| Hosea Kambonde[15] | IT Systems Developer |
| *Innocentia Mpofana[2] | Diagnostic Laboratory Manager |
| Jabu Kwinda[14] | Driver |
| *Jaco Dreyer[2] | Senior Research Data Manager |
| Jade Cousins[2] | Laboratory Technologist |
| Jaikrishna Kalideen[16] | Radiologist |
| *Janet Seeley[6] | Co-investigator |
| Kandaseelan Chetty[2] | Laboratory Technician |
| *Kathy Baisley[2,6] | Co-investigator |
| Kayleen Brien[2] | Laboratory Technologist |
| Kennedy Nyamande[17] | Pulmonology Consultant |
| Kgaugelo Moropane[14] | Radiographer |
| Khabonina Malomane[14] | Radiographer |
| *Khadija Khan[2] | Biorepository Manager |
| Khanyisani Buthelezi[2] | Professional Nurse |
| Kimeshree Perumal[2] | Laboratory Intern |
| *Kobus Herbst[2] | Co-investigator |
| Lindani Mthembu[2] | Information Technology Assistant |
| Logan Pillay[2] | Laboratory Technician |
| Mandisi Dlamini[2] | Enrolled Nurse |
| Mandlakayise Zikhali[2] | Clinical Research Assistant Supervisor |
| *Mark Siedner[2,10,11,12] | Co-Principal Investigator |

| | |
|---|---|
| Mbali Mbuyisa[2] | Enrolled Nurse |
| Mbuti Mofokeng[2] | Clinical Specimen Driver/Laboratory Assistant |
| Melusi Sibiya[2] | Enrolled Nurse |
| Mlungisi Dube[2] | Clinical Research Assistant |
| Mosa Suleman[17] | Pulmonology Consultant |
| Mpumelelo Steto[2] | Driver |
| Mzamo Buthelezi[2] | Clinical Research Assistant |
| Nagavelli Padayachi[2] | Laboratory Technologist |
| Nceba Gqaleni[2,18] | Public Engagement |
| *Ngcebo Mhlongo[2] | Study Physician |
| Nokukhanya Ntshakala[2] | Laboratory Technician |
| Nomathamsanqa Majozi[2] | Public Engagement |
| Nombuyiselo Zondi[2] | Professional Nurse |
| Nomfundo Luthuli[2] | Laboratory Technician |
| Nomfundo Ngema[2] | Laboratory Technician |
| Nompilo Buthelezi[2] | Training Coordinator |
| Nonceba Mfeka[2] | Clinical Research Assistant |
| Nondumiso Khuluse[2] | Biorepository Laboratory Technician |
| Nondumiso Mabaso[2] | Laboratory Intern |
| Nondumiso Zitha[2] | Biorepository Research Assistant |
| Nonhlanhla Mfekayi[2] | Clinical Research Assistant |
| Nonhlanhla Mzimela[2] | Enrolled Nurse |
| Nozipho Mbonambi[2] | Professional Nurse |
| Ntombiyenhlanhla Mkhwanazi[2] | Clinical Research Assistant |
| Ntombiyenkosi Ntombela[2] | Enrolled Nurse |
| *Olivier Koole[2,6] | Co-Principal Investigator (2017-2019) |
| Pamela Ramkalawon[2] | Laboratory Research Technician |
| Pfarelo Tshivase[17] | Driver |
| Phakamani Mkhwanazi[2] | Clinical Research Assistant |

| | |
|---|---|
| Philippa Mathews[2] | Clinical Governance |
| Phumelele Mthethwa[2] | Enrolled Nurse |
| Phumla Ngcobo[2] | Communications |
| Ramesh Jackpersad[19] | Radiologist |
| Raynold Zondo[2] | Operational Oversight |
| *Resign Gunda[2,4,5] | Programme Manager |
| Rochelle Singh[2] | Laboratory Technician |
| Rose Myeni[2] | Clinical Research Assistant |
| *Sanah Bucibo[2] | Lead Nurse |
| Sandile Mthembu[2] | Enrolled Nurse |
| *Sashen Moodley[2] | Microbiology Laboratory Supervisor |
| Sashin Harilall[2] | Grants Office |
| Senamile Makhari[2] | Biorepository Laboratory Technician |
| Seneme Mchunu[2] | Information Technology Assistant |
| Senzeni Mkhwanazi[2] | Clinical Research Assistant |
| Sibahle Gumbi[2] | Research Admin Assistant |
| Siboniso Nene[2] | Professional Nurse |
| Sibusiso Mhlongo[2] | Driver |
| Sibusiso Mkhwanazi[2] | Driver |
| Sibusiso Nsibande[2] | Driver |
| Simphiwe Ntshangase[2] | Laboratory Technician/LIMS Administrator |
| Siphephelo Dlamini[2] | AHRI Nursing Manager |
| Sithembile Ngcobo[2] | Laboratory Technologist |
| Siyabonga Nsibande[2] | General Worker |
| *Siyabonga Nxumalo[2] | Research Data Manager |
| Sizwe Ndlela[2] | Laboratory Technician |
| Skhumbuzo Mthombeni[2] | General Worker |
| Smangaliso Zulu[2] | Clinical Research Assistant |
| Sphiwe Clement Mthembu[2] | General Worker |

| | |
|---|---|
| Sphiwe Ntuli[2] | Professional Nurse |
| *Stephen Olivier[2] | Statistician |
| Talente Ntimbane[2] | Enrolled Nurse |
| Thabile Zondi[2] | Laboratory Technologist |
| *Thandeka Khoza[2] | Co-investigator (2019-present) |
| Thengokwakhe Nkosi[2] | Driver |
| *Theresa Smit[2] | Head: Diagnostic Research |
| Thokozani Bhengu[2] | Enrolled Nurse |
| Thokozani Simelane[2] | Professional Nurse |
| *Thumbi Ndung'u[2,5,7,8,9] | Co-investigator |
| *Tshwaraganang Modise[2] | Research Data Manager |
| Tumi Madolo[2] | Research Data Manager |
| Velile Vellem[14] | Driver |
| Welcome Petros Mthembu[2] | Enrolled Nurse |
| *Willem Hanekom[2] | Principal Investigator (2019-present) |
| Xolani Mkhize[2] | Enrolled Nurse |
| Zamashandu Mbatha[2] | Enrolled Nurse |
| Zinhle Buthelezi[2] | Enrolled Nurse |
| Zinhle Mthembu[2] | Enrolled Nurse |
| *Zizile Sikhosana[2] | Somkhele Laboratory Supervisor |
